# Electroconvulsive Therapy Drives Sensorimotor Network Segregation in Depression: A Multiscale Edge-Centric Connectomic Study

**DOI:** 10.64898/2026.07.29.26358453

**Authors:** Kexu Zhang, Jiang Long, Rui Li, Xiaochi Yuan, Chenchen Zhang, Ranran Xue, Liju Qian, Jiaojian Wang, Yanghua Tian, Wei Deng, Kun Li

**Affiliations:** Department of Physical Therapy, Shandong Daizhuang Hospital, Jining, 272075, China; Jining Key Laboratory of Neuromodulation, Jining, 272075, China; Mental Health Center, National Center for Mental Disorders, West China Hospital of Sichuan University, Chengdu, 610041, China; State Key Laboratory of Primate Biomedical Research, Institute of Primate Translational Medicine, Kunming University of Science and Technology, Kunming, 650500, China; The First Affiliated Hospital of USTC, University of Science and Technology of China, Hefei, 230000, China; Department of Neurology, the Second Affiliated Hospital of Anhui Medical University, Hefei, 230601, China; Affiliated Mental Health Center & Hangzhou Seventh People’s Hospital, Zhejiang University School of Medicine, Hangzhou, 310058, China; Liangzhu Laboratory, MOE Frontier Science Center for Brain Science and Brain-machine Integration, State Key Laboratory of Brain-machine Intelligence, Zhejiang University, Hangzhou, 311121, China

**Keywords:** Electroconvulsive therapy, Major depressive disorder, Edge-centric connectomics, sensorimotor network, Connectome entropy, Multiscale decoding

## Abstract

**Background:** Electroconvulsive therapy (ECT) induces widespread brain effects and remains the most effective intervention for severe major depressive disorder (MDD). However, how ECT reshapes the global organization of functional connectomes remains poorly understood. Edge-centric connectomics offers a framework for characterizing large-scale reconfiguration beyond conventional node-based analyses.

**Methods:** Longitudinal resting-state fMRI data from a primary cohort (80 MDD patients, 75 healthy controls) and an independent validation cohort (30 MDD patients) were analyzed. Edge-centric normalized entropy was utilized to quantify connectomic topology at baseline and post-ECT. These topological changes were evaluated for clinical associations and multiscale spatial correlations encompassing cognitive dimensions, neurotransmitter maps, and transcriptomic profiles. Additionally, baseline edge-centric features were leveraged in a machine learning framework to predict treatment response.

**Results:** At baseline, MDD patients showed increased entropy in the subcortical network and decreased entropy in the dorsal attention and sensorimotor networks. Following ECT, a further reduction in sensorimotor network (SMN) entropy was observed, which was replicated in the independent cohort. SMN reorganization was significantly associated with improvements in specific depressive symptoms. Multiscale decoding revealed that these topological shifts spatially aligned with broad monoaminergic receptor distributions and transcriptomic signatures governing neuroplasticity and specific cell types. Furthermore, baseline edge-centric features outperformed conventional fMRI metrics in predicting treatment response and maintained partial cross-site generalizability.

**Conclusions:** ECT is associated with selective reorganization of the sensorimotor network rather than normalization of baseline abnormalities. Edge-centric connectomics combined with multiscale biological annotations provides a robust framework for characterizing therapeutic mechanisms and developing predictive biomarkers in MDD.

**Clinical trial registration:** ChiCTR2400093114, Chinese Clinical Trial Registry (www.chictr.org.cn).

## 1 Introduction

Major depressive disorder (MDD) is one of the most prevalent and disabling psychiatric disorders worldwide, characterized by persistent affective, cognitive, and somatic symptoms that substantially impair quality of life and social functioning (1,2). Although antidepressant medications and psychotherapy are considered first-line treatments, a considerable proportion of patients fail to achieve adequate symptom remission, ultimately developing treatment-resistant depression (TRD) (3). For these refractory cases, as well as for patients presenting with acute suicidal risk, electroconvulsive therapy (ECT) remains the most rapidly acting and highly efficacious intervention available, achieving clinical response rates that significantly surpass conventional treatments (4,5). However, despite its unparalleled clinical efficacy, the broader utilization of ECT frequently encounters challenges such as cognitive adverse effects, historical stigma, and persistent public misconceptions (6). Importantly, the neurobiological mechanisms underlying its therapeutic effects remain insufficiently understood. This mechanistic gap not only perpetuates societal stigma surrounding ECT, but also constrains the development of mechanism-informed treatment optimization strategies and reliable biomarkers for patient stratification.

Over the past decades, multimodal neuroimaging has emerged as a pivotal tool for unraveling the macroscopic neural mechanisms of ECT, although the majority of these investigations have been constrained by relatively small sample sizes(7,8). Structural magnetic resonance imaging (MRI) studies have consistently demonstrated robust gray matter alterations following ECT, particularly within limbic and subcortical regions such as the hippocampus and amygdala (9,10). However, structural neuroimaging primarily characterizes localized regional effects and inherently lacks the sensitivity to capture the rapid, dynamic functional reorganization associated with clinical response (11). To address this limitation, resting-state functional MRI (rs-fMRI) has been widely applied to characterize ECT-related functional network alterations. Using traditional node-centric functional connectivity (FC) frameworks, prior studies have reported altered integration within canonical macro-scale circuits, such as increased connectivity in the prefrontal, default mode, and limbic networks (12,13), alongside reduced connectivity within hippocampus-thalamus-striatum pathways(14,15). However, these findings remain largely confined to simple pairwise interaction models. Given the profound, whole-brain modulatory effects of ECT, this reductionist framework inherently fails to capture higher-order topological reconfigurations. In physiological reality, true therapeutic mechanisms likely require the dynamic, simultaneous participation of individual brain hubs across multiple overlapping networks (16). Therefore, there is a critical need for analytic frameworks capable of capturing higher-order patterns of functional organization to better elucidate the systems-level mechanisms of ECT.

To overcome these limitations, emerging edge-centric functional connectivity frameworks have provided a fundamentally new perspective for characterizing large-scale brain organization (17). Rather than evaluating isolated interactions between brain regions, edge-centric analyses quantify the temporal co-fluctuation between the connections themselves, thereby capturing intricate network dynamics that remain inaccessible to conventional node-centric methods. Crucially, this framework naturally maps shared community architectures, directly addressing the biological necessity of hubs engaging concurrently across diverse functional systems. Such approaches have increasingly demonstrated their value in neuropsychiatric research, revealing abnormal higher-order network organization across disorders including schizophrenia (18), Alzheimer’s disease (19), autism spectrum disorder (20), and MDD (21–23). In particular, recent studies in MDD have identified disrupted edge-centric organization within prefrontal-striatal-thalamic circuits implicated in reward processing and cognitive control (21,22). Moreover, edge-centric connectivity patterns have shown substantial individual specificity and strong discriminative capacity between patients and healthy controls, highlighting their potential utility as biologically meaningful imaging biomarkers (23). Given their ability to capture disease-related network abnormalities in MDD, edge-centric approaches may also provide a valuable framework for characterizing the large-scale functional reorganization induced by ECT and for identifying biomarkers associated with treatment response.

In the present study, we applied an edge-centric connectomic framework to investigate large-scale functional network reorganization associated with ECT in MDD. Leveraging a longitudinal dual-center cohort of over 100 patients, we systematically characterized ECT-induced alterations in edge-centric organization at both macroscopic network and regional levels. To evaluate the translational potential of these edge-centric features, we developed predictive models to classify categorical treatment response and forecast the depressive symptom improvement. In addition, to provide biological interpretation of the observed topological changes, we integrated multiscale cross-modal analyses, linking edge-centric reorganization to cognitive functional annotations, neurotransmitter receptor distributions, and spatial transcriptomic expression profiles. Building upon this comprehensive framework, we hypothesized that ECT may exert its profound therapeutic effects through previously unrecognized mechanisms of higher-order, large-scale brain reorganization.

## 2 Methods and Materials

### 2.1 Participants

A total of 110 patients with MDD and 75 healthy controls (HCs) were enrolled in this study. The primary cohort was recruited at Shandong Daizhuang Hospital between December 2024 and January 2026, including 80 patients with MDD and 75 HCs (Site 1). Patients with MDD were diagnosed using the Structured Clinical Interview for DSM-5 Disorders (SCID-5) and met the following inclusion criteria: (1) age between 14 and 50 years; (2) right-handed Han Chinese ethnicity; (3) a 24-item Hamilton Depression Rating Scale (HAMD-24) score >20; (4) an intelligence quotient >90; and (5) scheduled to receive ECT. Exclusion criteria included substance dependence, other psychiatric disorders, traumatic brain injury, epilepsy or other organic brain diseases, severe physical or metabolic disorders, hepatic or renal dysfunction, contraindications to MRI or ECT, pregnancy or breastfeeding, a personal or family history of epilepsy, and receipt of other physical treatments during ECT. HCs were recruited from the local community and screened using the SCID-5. They were right-handed Han Chinese individuals aged 14–50 years, with an intelligence quotient >90 and no current or lifetime psychiatric disorders, major physical illnesses, neurological disorders, MRI contraindications, or use of medications affecting the central nervous system. An independent validation cohort consisting of 30 patients with MDD was recruited from Anhui Mental Health Center (Site 2). Detailed inclusion and exclusion criteria for the validation cohort are provided in the Supplementary Materials. Demographic and clinical characteristics of all participants are summarized in Table 1. Written informed consent was obtained from all participants or their legal guardians prior to study enrollment. This study was approved by the Ethics Committee of Shandong Daizhuang Hospital (Approval No. 2023KY022-202312KS-2) and was registered at Chinese Clinical Trial Registry (Registration No. ChiCTR2400093114).

**Table 1.** Demographic and clinical characteristics of the study participants.

| Characteristic | HC (Site 1) | MDD (Site 1) | MDD (Site 2) |
| --- | --- | --- | --- |
| Number of subjects | 75 | 80 | 30 |
| Sex (Male/Female) | 33/42 | 27/53 | 6/24 |
| Age (years) | 19.40±3.73 | 20.70±7.16 | 33.77±8.91 |
| Duration of illness (months) | / | 44.91±36.71 | 56.23±68.80 |
| HAMD scores<sup>a</sup> | / | 35.40±6.93 | 24.30±5.95 |
| HAMA scores | / | 27.78±10.05 | 22.73±6.70 |

*Note:* Data are presented as mean ± standard deviation (SD) or N/N. MDD, major depressive disorder; HC, healthy controls; HAMD, Hamilton Depression Rating Scale; HAMA, Hamilton Anxiety Rating Scale. <sup>a</sup>MDD in Site 1 was assessed using the 24-item HAMD, whereas MDD in Site 2 was assessed using the 17-item HAMD.

### 2.2 ECT Procedure

ECT was administered using a Thymatron System IV device (Somatics, LLC). Prior to each stimulation, general anesthesia was induced with intravenous propofol (1.0-1.5 mg/kg), and neuromuscular blockade was achieved using succinylcholine (1.0 mg/kg). Atropine (0.5 mg) was additionally administered to inhibit glandular secretions and prevent bradycardia. For patients in the Site 1, standard bitemporal electrode placement was utilized. The initial stimulus intensity was determined using an age-based dosing method (e.g., 45 years, 45% of maximum output). If an adequate seizure was not successfully induced, the stimulus energy was incrementally increased by 5% during the same session until a generalized seizure was confirmed both visually and electroencephalographically (EEG). ECT sessions were conducted every other day (typically three times per week). The total number of treatment sessions and the endpoint of the ECT course were individually determined by experienced psychiatrists, based on the patient’s clinical improvement and tolerability. For patients in the external validation cohort, bifrontal electrode placement was applied.

### 2.3 Clinical Assessments

In Site 1, the severity of baseline and post-treatment depressive symptoms was evaluated for all MDD patients using the 24-item HAMD (24). To further delineate the multidimensional therapeutic effects of ECT, the HAMD score was decomposed into five distinct symptom dimensions: psychic depression (items 1, 2, 3, 8, 22, 23, and 24), loss of motivated behavior (items 7, 12, 14, and 16), disturbed thinking (items 17, 19, 20, and 21), anxiety (items 9, 10, 11, and 15), and sleep disturbance (items 4, 5, and 6) (25). General anxiety severity was independently measured using the Hamilton Anxiety Rating Scale (HAMA) (26). Furthermore, given the well-documented rapid anti-suicidal efficacy of ECT, changes in the suicidal ideation severity were specifically quantified using the subscale of the Columbia-Suicide Severity Rating Scale (C-SSRS) (27). For Site 2, longitudinal changes in depressive severity before and after ECT were monitored using the 17-item HAMD.

### 2.4 Edge-Centric Community Analysis

High-resolution structural and resting-state functional MRI data were acquired for all participants at both Site 1 and Site 2 before and after ECT treatment. Detailed MRI acquisition parameters and standard image preprocessing procedures are provided in the Supplementary Materials.

The analysis pipeline for edge-centric Community is illustrated in Figure 1A (17). Following preprocessing, the whole brain was parcellated into 246 regions of interest (ROIs) using the Brainnetome Atlas (28), and the mean blood-oxygen-level-dependent (BOLD) time series were extracted for each ROI. To construct the edge-centric network, the regional time series were first z-scored. Subsequently, the edge time series for each pair of ROIs were generated by calculating the element-wise product of their respective z-scored regional time series. This approach unfolds the conventional static functional connectivity into dynamic co-fluctuation signals at a single-frame resolution. Finally, the edge functional connectivity (eFC) matrix was constructed by computing the Pearson correlation coefficients between all pairs of edge time series.

**Figure 1.**
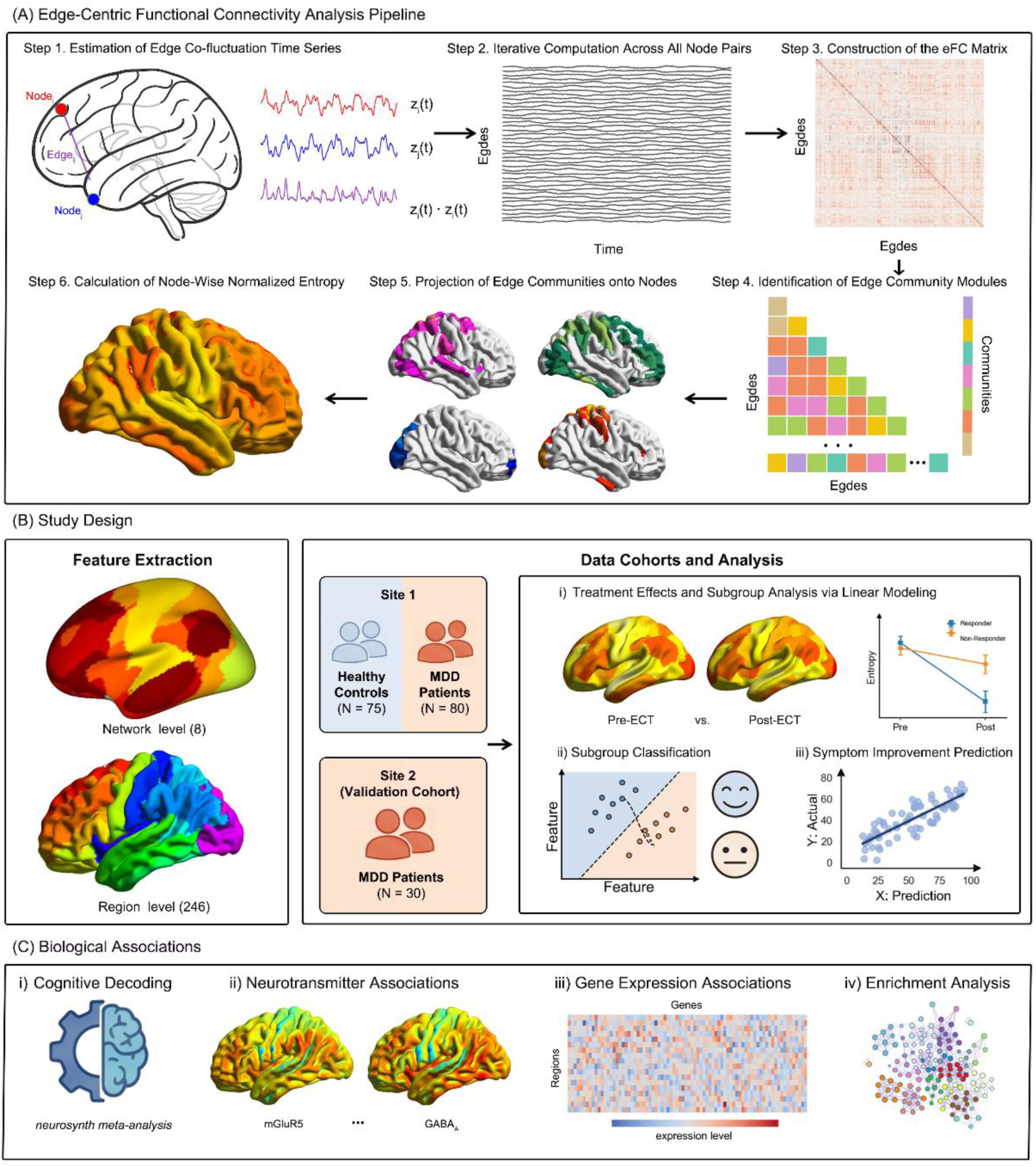
Study design and analytical framework. (A) Pipeline for edge-centric functional connectivity (eFC) analysis. Edge co-fluctuation time series are estimated from resting-state fMRI to capture edge-level dynamics, enabling eFC matrix construction, module identification, and the derivation of node-wise normalized entropy. (B) Cohorts and analytical workflow. Features were extracted at macroscopic (8 networks) and mesoscopic (246 regions) scales from 110 MDD patients and 75 healthy controls across a discovery (Site 1) and an independent validation (Site 2) cohort. Analyses include linear modeling of ECT treatment effects, machine learning classification of treatment responsivity, and regression prediction of symptom improvement. (C) Multimodal biological associations. Significant ECT-induced eFC alterations were linked to underlying biological mechanisms via spatial correlation with Neurosynth cognitive meta-analyses, neurotransmitter receptor/transporter distributions, and AHBA transcriptomic profiles, followed by pathway and cell-type enrichment.

Given the substantial computational challenges posed by the high dimensionality of the eFC matrix, we applied a low-dimensional clustering approach to detect overlapping network communities. First, the eFC matrix underwent eigenvector decomposition to reduce dimensionality, and the rescaled edge features were partitioned into non-overlapping clusters using the k-means algorithm. Next, to characterize the functional profile of individual brain regions, the edge-based community labels were projected back onto their corresponding nodes. Because each node participates in multiple edges, this projection yields a multidimensional overlapping community pattern for every specific region. To quantify this functional diversity, we calculated the normalized entropy of the community assignment distribution for each node. The regional normalized entropy serves as a dynamic index ranging from 0 to 1, where higher values indicate that a region’s connections are uniformly distributed across diverse functional communities (reflecting high functional integration), whereas lower values suggest isolated, single-network affiliations. Detailed mathematical algorithms are provided in the Supplementary Materials.

### 2.5 Statistic Analysis

All statistical analyses were conducted using the R statistical programming environment and JAMOVI software (Version 2.6). The primary statistical inferences were based on data from Site 1. To investigate baseline differences in normalized entropy between the MDD and healthy control groups, general linear models were employed, incorporating age and sex as nuisance covariates. To precisely assess longitudinal alterations in normalized entropy induced by ECT within the MDD group, linear mixed-effects models were utilized. In these models, age, sex, illness duration, and the number of ECT sessions were entered as fixed effects, while subject identity was modeled as a random intercept to account for intra-subject correlations across repeated measures. Furthermore, associations between normalized entropy and clinical symptom severity were evaluated using partial Pearson’s or Spearman’s rank correlation coefficients. Comparative analyses of normalized entropy were conducted at two distinct topological scales: the network level and the region level. The network level analysis involved eight macroscopic networks, including the seven canonical cortical networks defined by Yeo’s functional parcellation: Visual (VIS), sensorimotor (SMN), Dorsal Attention (DAN), Ventral Attention (VAN), Limbic (LIM), Frontoparietal (FPN), and Default Mode (DMN), as well as an additional Subcortical Network (SCN) (29). Region-level analyses were based on the 246 specific regions of interest (ROIs) from the Brainnetome Atlas, with each ROI systematically assigned to one of the aforementioned eight large-scale networks based on maximum spatial overlap (28). Furthermore, the MDD cohort was stratified into responder and non-responder subgroups based on a clinical response threshold defined as a ≥ 50% reduction in the HAMD score, and all relevant statistical analyses were subsequently repeated within these subgroups. Finally, to strictly control the false positive rate arising from multiple testing across nodes and networks, all statistical p-values were adjusted using the false discovery rate (FDR) correction procedure.

### 2.6 Independent Validation and Predictive Modeling

To assess the reproducibility of our findings, all aforementioned statistical analyses were strictly replicated using the independent validation cohort (Site 2), and the results were systematically compared with those from Site 1.

Furthermore, to evaluate the clinical utility of network dynamics, predictive models were developed utilizing baseline normalized entropy combined with demographic and clinical features. The prediction framework encompassed two complementary tasks: a classification model to differentiate ECT responders from non-responders, and a regression model to predict the continuous reduction rate of HAMD scores. To rigorously evaluate model performance and generalizability, two validation strategies were implemented: an internal leave-one-out cross-validation (LOOCV) within Site 1, and an external independent validation where the models were trained exclusively on Site 1 and tested on Site 2. Moreover, to benchmark the prognostic value of normalized entropy against conventional resting-state fMRI metrics, we introduced the fractional amplitude of low-frequency fluctuations (fALFF) as an index of local regional activity, and degree centrality (DC) as a measure of nodal functional connectivity. The predictive performance was systematically compared across four distinct feature configurations: clinical features alone, clinical features combined with normalized entropy, clinical features combined with fALFF, and clinical features combined with DC. Detailed methodological procedures regarding the machine learning algorithms and predictive modeling are provided in the Supplementary Materials.

### 2.7 Multiscale Biological and Cognitive Annotation of Network Reorganization

To uncover the multi-scale mechanisms underlying ECT-induced network reorganization, we performed spatial association analyses linking regional entropy changes to macro-scale cognition and micro-scale biological architectures. First, the spatial maps of entropy changes were correlated with topic-based meta-analytic maps from the Neurosynth database to decode the implicated cognitive and behavioral domains. Next, to bridge these topological reconfigurations with neurochemical systems, the regional entropy alterations were spatially correlated against normative whole-brain density maps of diverse neurotransmitter receptors and transporters. Finally, at the molecular scale, we mapped the macroscopic entropy alterations onto regional transcriptomic profiles derived from the Allen Human Brain Atlas (AHBA). Genes whose spatial expression patterns exhibited significant associations with the entropy changes were subsequently subjected to functional pathway and cell-type-specific enrichment analyses. Detailed procedures for these cross-scale mappings, permutation tests, and enrichment analyses are provided in the Supplementary Materials.

## 3 Results

### 3.1 Baseline Alterations of Edge-Centric Normalized Entropy in MDD

Baseline differences in edge-centric community entropy were first examined between healthy controls (n = 75) and patients with MDD (n = 80) from Site 1. No significant between-group differences were observed in age (t = 1.404, p = 0.162) or sex distribution (χ² = 1.714, p = 0.190).

To characterize large-scale network alterations, linear regression analyses were performed with group as the effect of interest while controlling for age and sex. Comparisons of normalized entropy across the eight canonical functional networks revealed significantly increased entropy within the SCN in patients with MDD relative to HCs (standardized β = 0.639, t = 4.123, p < 0.001, FDR-corrected p < 0.001). In contrast, significantly reduced entropy was observed in the DAN (standardized β = - 0.479, t = -3.001, p = 0.003, FDR-corrected p = 0.012). A trend-level decrease was additionally identified in the SMN (standardized β = -0.376, t = -2.328, p = 0.021, FDR-corrected p = 0.056). No significant alterations were detected in the remaining functional networks (Figure 2A).

**Figure 2.**
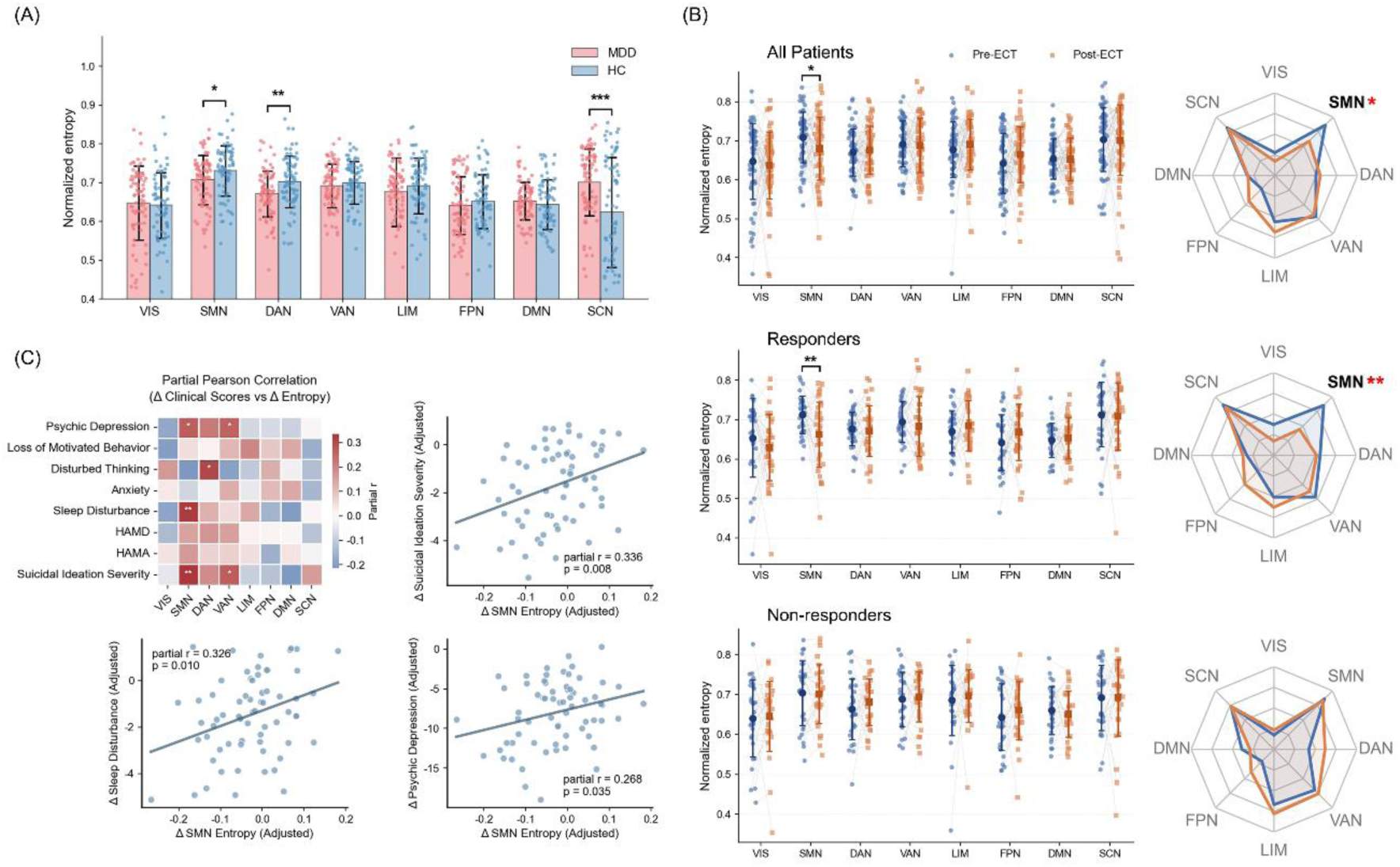
Baseline alterations and longitudinal ECT effects on edge-centric network entropy at Site 1. (A) Baseline comparison of normalized entropy between MDD patients and healthy controls (HC) across 8 canonical networks. MDD patients exhibit significantly decreased entropy in the SMN and DAN, alongside significantly increased entropy in the SCN. (B) Longitudinal changes in normalized entropy from pre-ECT to post-ECT. At the whole-group level (top), SMN entropy significantly decreases following treatment. Subgroup analyses reveal that this significant reduction in SMN entropy is specifically driven by the Responder group (middle), whereas no significant changes are observed in the Non-responder group (bottom). Radar charts (right) summarize the mean network entropy shifts across groups. (C) Partial Pearson correlation analysis between ECT-induced changes in normalized entropy and changes in clinical symptom scores. The heatmap (left) displays the correlation matrix across multiple symptom domains. Scatter plots (right) illustrate representative significant associations, demonstrating that greater reductions in SMN entropy correlate with greater improvements in suicidal ideation severity, sleep disturbance, and psychic depression. *p < 0.05, p < 0.01, p < 0.001. MDD, major depressive disorder; HC, healthy controls; ECT, electroconvulsive therapy; HAMD, Hamilton Depression Rating Scale; HAMA, Hamilton Anxiety Rating Scale; VIS, visual network; SMN, sensorimotor network; DAN, dorsal attention network; VAN, ventral attention network; LIM, limbic network; FPN, frontoparietal network; DMN, default mode network; SCN, subcortical network.

At the regional level, significant entropy abnormalities were predominantly localized to subcortical and paralimbic regions, including the thalamus, basal ganglia, and cingulate cortex (Table S2). Associations between baseline symptom severity and regional normalized entropy are presented in the Supplementary Materials.

### 3.2 Longitudinal Alterations of Edge-Centric Normalized Entropy Following ECT

Due to early discharge, refusal to complete follow-up MRI scanning, or excessive head motion, a subset of patients from Site 1 was excluded from the longitudinal analyses. The final paired pre- and post-treatment dataset consisted of 66 patients with MDD. Following ECT treatment, significant reductions were observed in overall depressive severity, anxiety symptoms, suicidal ideation severity, and all symptom subscales derived from the HAMD (all p < 0.001). Longitudinal changes in network-level normalized entropy were evaluated using linear mixed-effects models. Among the eight large-scale functional networks, a trend-level longitudinal reduction was identified in the SMN (t = 2.588, p = 0.012, FDR-corrected p = 0.096; Figure 2B), indicating reduced overlapping community entropy following ECT.

Based on treatment response criteria, 36 patients were classified as responders and 30 as non-responders, corresponding to an overall response rate of 54.6% (Table S1). Longitudinal symptom changes for both groups are illustrated in Figure S2. Subgroup analyses demonstrated that the decrease in SMN entropy was primarily driven by the responder group. Specifically, responders showed a significant post-treatment reduction in SMN entropy (t = 3.269, p = 0.003, FDR-corrected p = 0.024), whereas no significant change was observed in non-responders (t = 0.186, p = 0.854).

Furthermore, partial correlation analyses indicated that the reduction in SMN entropy was significantly associated with specific clinical improvements, notably correlating with decreased suicidal ideation severity (partial r = 0.336, p = 0.008), Psychic Depression subscale scores (partial r = 0.268, p = 0.035), and Sleep Disturbance subscale scores (partial r = 0.326, p = 0.010) (Figure 2C).

At the regional level, the full cohort exhibited uncorrected entropy decreases in the postcentral gyrus and superior frontal gyrus, alongside increases in the hippocampus; however, none survived intra-network FDR correction (Figure 3, Table S3). In contrast, the responder subgroup demonstrated a significant entropy decrease in the postcentral gyrus and an increase in the inferior parietal lobule, both robustly surviving intra-network FDR correction. No significant regional network alterations were observed in non-responders (Figure S3, Tables S4-S5).

**Figure 3.**
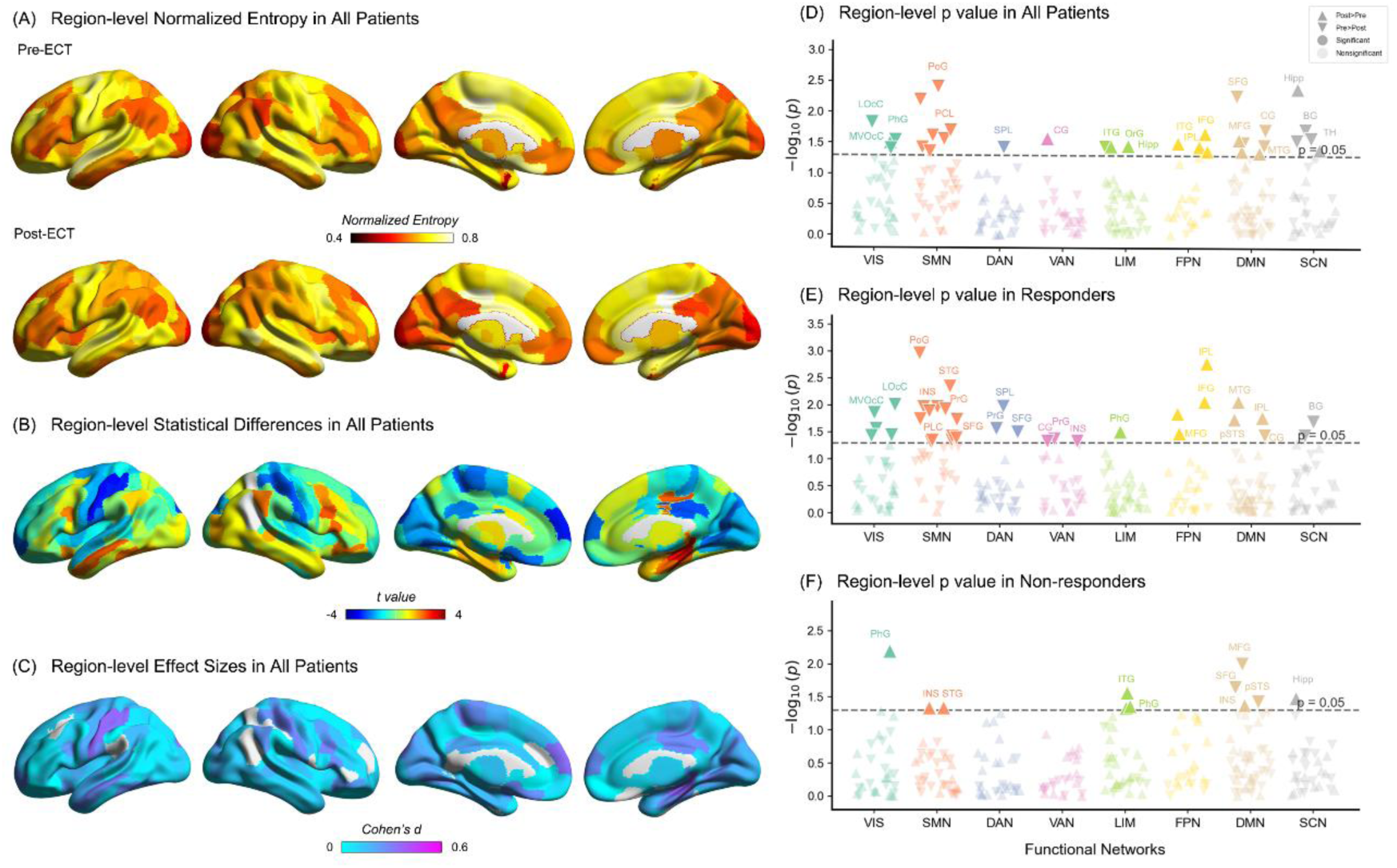
Region-level longitudinal alterations in edge-centric network entropy induced by ECT. (A) Spatial distribution of region-level normalized entropy in all patients before (Pre-ECT) and after (Post-ECT) treatment. (B, C) Brain surface maps depicting the unthresholded statistical differences (t-values) (B) and corresponding effect sizes (Cohen’s d) (C) of ECT-induced entropy changes across all patients. (D–F) Manhattan plots displaying the region-specific statistical significance of longitudinal entropy changes in (D) all patients, (E) the Responder subgroup, and (F) the Non-responder subgroup. Brain regions are grouped along the x-axis according to their canonical functional networks. VIS, visual network; SMN, sensorimotor network; DAN, dorsal attention network; VAN, ventral attention network; LIM, limbic network; FPN, frontoparietal network; DMN, default mode network; SCN, subcortical network.

### 3.3 Cross-Cohort Validation and Spatial Consistency of Edge-Centric Network Remodeling

To validate the reproducibility of our findings, longitudinal changes in normalized entropy were further examined in the validation cohort from Site 2 (n = 30). Using a linear mixed-effects model, we evaluated the treatment-related effects across the eight canonical functional networks (Figure 4A). Consistent with the discovery cohort, ECT induced a trend-level longitudinal reduction in SMN entropy (t = 2.595, p = 0.015, FDR-corrected p = 0.060). Moreover, a highly significant entropy decrease was identified in the subcortical network (SCN) (t = 3.687, p < 0.001, FDR-corrected p = 0.007). These network-level trajectories were reliably preserved in the responder subgroup (n = 23), which exhibited consistent post-treatment entropy reductions in both the SMN (t = 2.672, p = 0.014, FDR-corrected p = 0.056) and the SCN (t = 4.619, p < 0.001, FDR-corrected p < 0.001).

**Figure 4.**
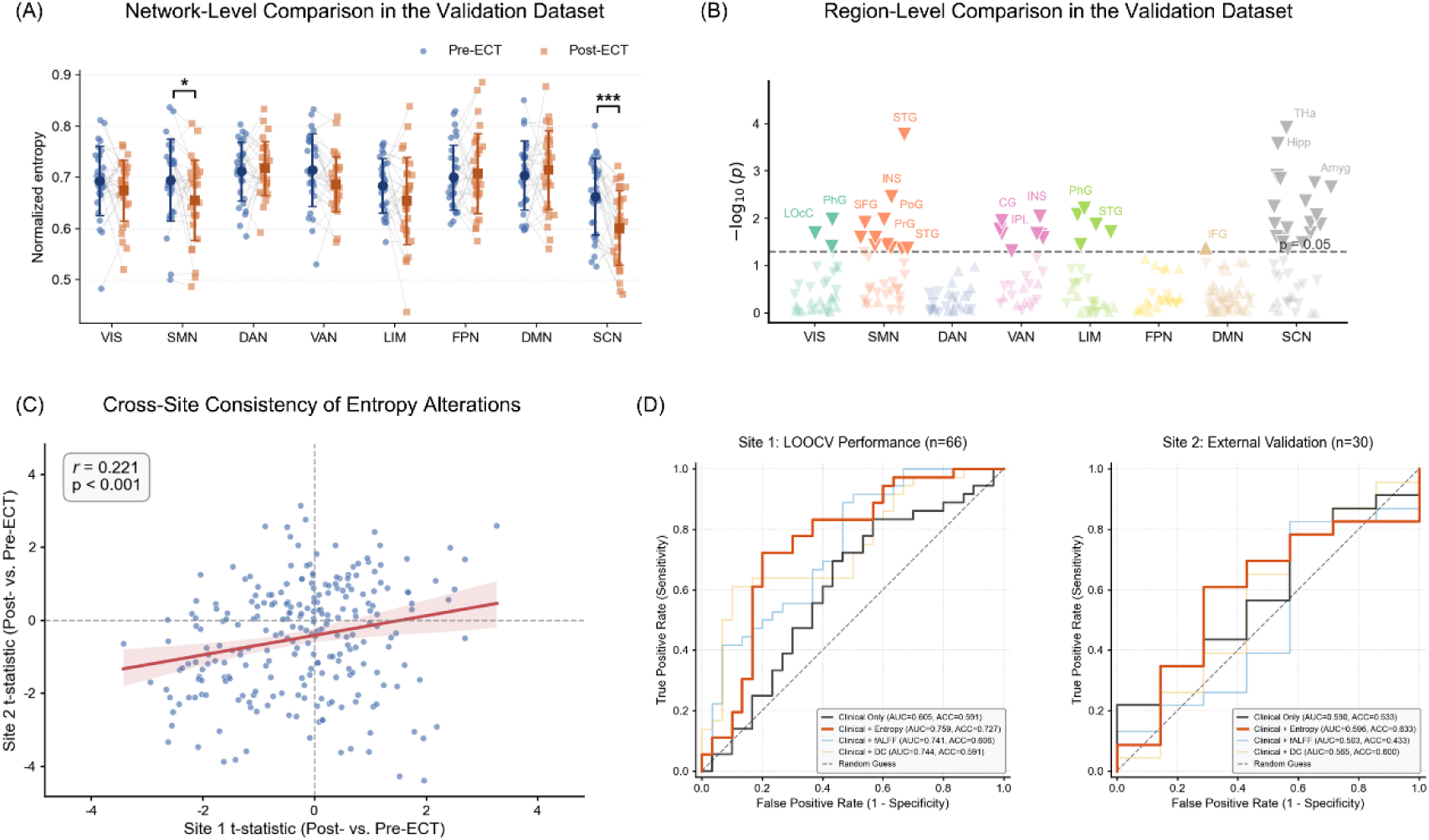
External validation and predictive modeling of ECT efficacy using edge-centric network entropy. (A) Network-level longitudinal comparison of normalized entropy in the independent validation dataset (Site 2). Consistent with the discovery cohort, the SMN exhibited a significant decrease following ECT; additionally, a significant reduction in the SCN entropy was also observed in this dataset. (B) Manhattan plot illustrating the region-level statistical significance of longitudinal entropy changes in the validation dataset. (C) Cross-site consistency of ECT-induced entropy alterations. A significant positive correlation (r = 0.221, p < 0.001) was observed between the region-level t-statistic maps (Post- vs. Pre-ECT) of Site 1 and Site 2, demonstrating high spatial reproducibility of the treatment effects. (D) Machine learning classification performance for predicting ECT efficacy (Responder vs. Non-responder). ROC curves are shown for the LOOCV within Site 1 (left) and the external validation in Site 2 (model trained on Site 1, right). The model combining pre-ECT normalized entropy with clinical information achieved the highest predictive performance, outperforming models using clinical information alone or combined with conventional neuroimaging features. *p < 0.05, ***p < 0.001. ECT, electroconvulsive therapy; ROC, receiver operating characteristic; AUC, area under the curve; LOOCV, leave-one-out cross-validation; fALFF, fractional amplitude of low-frequency fluctuation; DC, degree centrality; VIS, visual network; SMN, sensorimotor network; DAN, dorsal attention network; VAN, ventral attention network; LIM, limbic network; FPN, frontoparietal network; DMN, default mode network; SCN, subcortical network.

At the regional level, post-treatment entropy differences were predominantly localized to the thalamus, superior temporal gyrus, and hippocampus (Figure 4B, Table S6). To quantitatively assess the cross-cohort spatial consistency of these regional alterations, we conducted spatial correlation analyses of the region-level t-statistics between Site 1 and Site 2. Although no significant spatial correlation was detected across the full samples (p > 0.05), a robust inter-site spatial correlation of regional entropy changes was identified specifically within the responder subgroups (r = 0.221, p < 0.001, Figure 4C). This robust cross-cohort spatial correspondence further underscores a replicable, neurobiologically specific pattern of network remodeling associated with ECT.

### 3.4 Predictive Performance of Edge-Centric Normalized Entropy for ECT Treatment Response

Classification and regression models were developed to evaluate the translational potential of edge-centric features in predicting ECT treatment response (Table S7, Figure S4). Within the internal cross-validation framework (Site 1), the model integrating normalized edge-centric entropy with clinical variables (Clin + ENORM) achieved the highest classification performance (area under the curve [AUC] = 0.759, accuracy = 72.7%). This combined model substantially outperformed the clinical-only baseline (AUC = 0.605, accuracy = 59.1%), as well as conventional node-centric neuroimaging baselines, including Clin + fALFF (AUC = 0.741, accuracy = 60.6%) and Clin + DC (AUC = 0.744, accuracy = 59.1%). During independent external validation (Site 2), despite an anticipated cross-site attenuation in predictive performance, the Clin + ENORM model successfully maintained the highest generalization accuracy (63.33%) (Figure 4D).

Furthermore, in regression analyses predicting continuous HAMD reduction rates, Clin + ENORM demonstrated the strongest internal correlation between predicted and observed improvement (r = 0.426, p < 0.001), outperforming the clinical-only model (r = 0.350, p = 0.004) and all conventional neuroimaging baselines. However, this regression efficacy was confined to the discovery cohort and did not generalize to the external validation set.

### 3.5 Multiscale Biological and Cognitive Annotation of ECT-Induced Edge-Centric Entropy Changes

To further elucidate the biological substrates of ECT-related network reorganization, we performed a multiscale spatial association analysis linking longitudinal changes in edge-centric normalized entropy with transcriptomic, neurochemical, and cognitive decoding maps (Figure 5A–E; Tables S8–S11).

**Figure 5.**
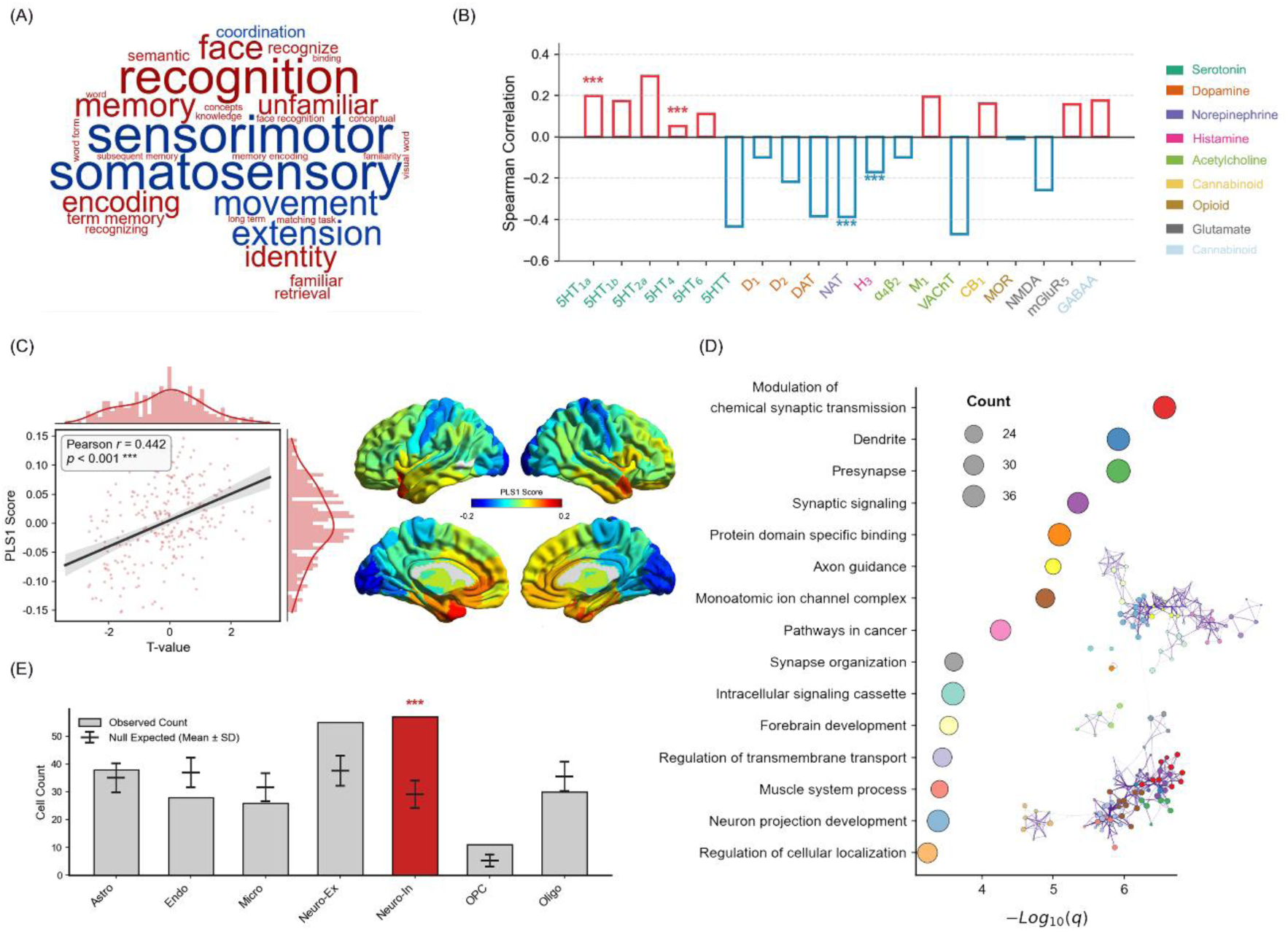
Multimodal biological mechanisms underlying ECT-induced edge-centric network alterations. (A) Cognitive decoding via Neurosynth meta-analysis. The word cloud displays the top 30 cognitive terms exhibiting the highest correlation weights with the ECT-induced entropy alterations. (B) Spatial Spearman correlations between the statistics of regional entropy alterations and the cortical distributions of diverse neurotransmitter receptors and transporters. (C) Transcriptomic associations using PLS regression. The scatter plot (left) reveals a significant positive correlation (Pearson r = 0.442, p < 0.001) between the PLS1 scores and ECT-induced entropy changes (t-values) at Site 1. The brain surface map (right) illustrates the spatial distribution of PLS1 weights, which identified a robust transcriptional signature of 620 significant genes. (D) Functional enrichment analysis of the 620 significant genes. Dot size indicates the gene count, and the x-axis represents the statistical significance. (E) Cell-type specific enrichment analysis. The bar chart compares the observed gene counts (bars) against the null expected background across distinct brain cell types, revealing a highly significant enrichment specifically in inhibitory neurons (Neuro-In). ***p < 0.001. PLS, partial least squares; Astro, astrocytes; Endo, endothelial cells; Micro, microglia; Neuro-Ex, excitatory neurons; OPC, oligodendrocyte precursor cells; Oligo, oligodendrocytes.

Meta-analytic decoding based on Neurosynth revealed that entropy alterations were predominantly negatively associated with sensorimotor processes, whereas positive associations were enriched in higher-order cognitive domains, particularly memory, recognition, and semantic processing (Figure 5A; Table S8).

At the chemoarchitectonic level, spatial correlation analyses revealed that the macro-scale network dynamics were significantly and specifically coupled with the distribution profiles of multiple neuromodulatory systems. Specifically, regional entropy changes exhibited robust positive spatial associations with serotonin receptors, including 5-HT_1A_ (ρ = 0.285, FDR-corrected p < 0.001) and 5-HT_4_ (ρ = 0.307, FDR-corrected p < 0.001). Conversely, significant negative correlations were identified with the norepinephrine transporter (NAT) (ρ = -0.264, FDR-corrected p < 0.001) and the histamine H3 receptor (ρ = -0.231, FDR-corrected p = 0.001) (Figure 5B, Figure S5, Table S9).

At the transcriptomic level, PLS analysis identified a dominant latent component (PLS1), which explained 19.52% of the covariance between gene expression and entropy alterations (Figure 5C). A total of 620 genes survived the threshold of FDR-corrected p < 0.001. Gene set enrichment analysis revealed that these genes were primarily enriched in synaptic signaling, neuronal structure organization, and neurodevelopmental pathways, particularly involving synaptic transmission, axon guidance, and dendritic function (Figure 5E; Table S10). Crucially, cell-type specific enrichment analysis demonstrated that this macro-scale transcriptomic signature was disproportionately overrepresented in inhibitory neurons (Figure 5D; Table S11).

## 4 Discussion

In this study, we applied an edge-centric connectomic framework to characterize large-scale functional reorganization associated with ECT in MDD. By quantifying the temporal co-fluctuations between connections rather than isolated regional activities, this approach transcends rigid nodal boundaries to precisely map the brain’s overlapping community architectures and high-order topological organization. Utilizing a robust dual-center cohort, we identified widespread abnormalities in edge-centric entropy in patients with MDD and demonstrated that ECT induced reproducible alterations in higher-order network architecture, particularly among treatment responders. Importantly, these network alterations were linked to clinical improvement, exhibited predictive utility for treatment outcomes, and converged across cognitive, neurochemical, and transcriptomic levels of analysis. Together, our findings support a model in which the antidepressant effects of ECT arise not simply from modulation of individual brain regions or pairwise connections, but from large-scale reorganization of overlapping functional communities across the brain.

Prior to examining the longitudinal effects of ECT, characterizing the baseline topological architecture confirms the reproducibility of the edge-centric framework in mapping MDD pathology. Compared to HCs, patients with MDD exhibited significantly elevated normalized entropy within the SCN. This finding is highly congruent with two large-scale consortium studies from the REST-meta-MDD project (N = 400 and N = 838) that utilized the identical analytical framework, consistently identifying subcortical hyper-connectivity, particularly in the thalamus, as a robust macroscopic signature of depression (21,22). From an edge-centric perspective, elevated entropy reflects a more distributed pattern of community participation, suggesting broader engagement of subcortical regions across multiple functional systems in MDD (30,31). In contrast, our analysis uniquely detected significantly reduced entropy in the DAN and SMN. Although alterations in these cortical networks were less prominent in previous large-scale studies employing the same methodology, this discrepancy may arise from at least two factors. First, the inherent variance within massive multi-center datasets, such as heterogeneous imaging protocols and site-specific noise, may attenuate some topological signatures (32). Second, clinical heterogeneity may contribute to these discrepant findings. Unlike previous studies that enrolled patients across a broad spectrum of illness severity, our cohort consisted exclusively of individuals referred for ECT, representing a clinically severe and largely treatment-resistant population. Within this context, the marked reductions in DAN and SMN entropy may reflect a more advanced stage of network dysfunction, characterized by diminished cross-community participation of systems involved in attentional control and sensorimotor processing (33,34).

Among all large-scale systems, the SMN emerged as a robust and reproducible target of ECT-induced network reorganization. Reduced SMN entropy was consistently observed in the primary cohort, with a stronger effect in treatment responders, and was independently replicated in the validation cohort. Although alterations within sensorimotor regions have been repeatedly reported in previous ECT studies, these findings have often been interpreted as secondary observations relative to changes in prefrontal and limbic circuitry (13,35–37). Our results suggest that the SMN may represent a central, yet underappreciated, substrate of therapeutic response. Importantly, the regions driving this effect extended beyond classical motor cortices and included the postcentral gyrus, superior temporal gyrus, and insula. Collectively, these regions support not only sensorimotor processing, but also auditory perception, interoceptive awareness, and the representation of bodily states (38,39). This pattern may reflect a broader sensorimotor–interoceptive network involved in integrating bodily and environmental information. Such a framework is particularly relevant to severe depression, which is frequently characterized by psychomotor retardation, altered bodily awareness, fatigue, and sleep disturbances (40,41). Notably, reduced SMN entropy was associated with greater improvements in depressive symptoms, suicidal ideation, and sleep quality. Within the edge-centric framework, lower entropy indicates reduced cross-community participation and a shift toward more functionally specialized network organization (42). Contrary to a normalization account, our findings suggest that ECT may further enhance the specialized organization of the SMN. This may strengthen the functional identity of sensorimotor and interoceptive systems, reducing their interaction with higher-order associative networks and supporting more stable processing of bodily and sensory information, ultimately contributing to clinical improvement (43,44). More broadly, these findings suggest that the therapeutic effects of ECT extend beyond affective and cognitive circuits. By reorganizing systems involved in bodily awareness, sensorimotor integration, and environmental engagement, ECT may act through partially embodied mechanisms. This may help explain the rapid improvement of psychomotor symptoms, suicidality, and sleep disturbance during treatment (45). However, the present findings likely capture network dynamics during the acute treatment phase. While increased specialization may support rapid symptom improvement, longer-term remission could involve additional reorganization of large-scale network interactions. Future longitudinal studies are required to characterize the temporal evolution of these effects.

To elucidate the microscale biological substrates driving this macroscopic topological reorganization, our multiscale spatial annotations revealed a highly convergent molecular landscape. At the neurochemical level, ECT-induced edge-centric alterations spatially co-segregated with the distribution of specific serotonergic receptors, most notably 5-HT1A and 5-HT4, thereby bridging our macroscopic network signatures with classical antidepressant pharmacological pathways(46). Conversely, significant negative spatial correlations were observed with the noradrenaline transporter and histamine H3 receptor distributions. Given the established roles of these systems in arousal, psychomotor regulation, and sleep–wake function (47,48), these findings suggest that ECT-related network reorganization may preferentially involve neurochemical architectures relevant to the improvement of psychomotor and sleep symptoms. Furthermore, our transcriptomic decoding revealed that regional entropy reconfigurations were tightly coupled with the expression profiles of genes governing synaptic signaling, axon guidance, and dendritic morphology, a physiological prerequisite for gating large-scale network co-fluctuations (49). Crucially, cell-type-specific enrichment localized these neuroplastic signatures predominantly to inhibitory neurons. Within cortical microcircuits, inhibitory interneurons act as the principal orchestrators of the excitation/inhibition (E/I) balance, a physiological prerequisite for gating large-scale network co-fluctuations (50). Overall, these findings converge to suggest a coordinated biological basis for ECT-induced network reorganization across molecular and cellular levels.

Beyond mechanistic insights, our findings also highlight the translational potential of edge-centric network organization for predicting ECT outcomes. Previous studies have demonstrated the utility of edge-centric frameworks in classifying psychiatric disorders, including autism, anxiety disorders, and MDD(23,51,52). Extending these findings, baseline edge-centric entropy improved the prediction of treatment response beyond conventional rs-fMRI metrics such as fALFF and DC, likely reflecting its ability to capture higher-order network organization that is inaccessible to regional activity or pairwise connectivity measures alone (53). Notably, although predictive performance decreased during external validation, the edge-centric model consistently achieved the highest accuracy for classifying responders across sites, suggesting a degree of robustness and generalizability. In contrast, prediction of continuous symptom improvement failed to generalize to the independent cohort, potentially reflecting inter-site differences in clinical characteristics and symptom distributions. Future studies with larger multi-center samples are warranted to further evaluate the clinical utility of edge-centric biomarkers for individualized treatment prediction.

Several limitations should be acknowledged. First, although our principal findings were replicated in an independent cohort, the relatively modest sample size of the validation dataset may have limited statistical power and the assessment of model generalizability. Second, because all patients received ECT, the absence of a medication-treated control group precludes determination of whether the observed network reorganization is specific to ECT or reflects more general mechanisms of symptom improvement. Finally, the transcriptomic and neurotransmitter analyses were based on normative datasets from healthy individuals rather than the studied patients and should therefore be interpreted as indirect biological inferences. Future studies incorporating patient-specific molecular data are needed to further validate these findings.

In conclusion, by integrating edge-centric network topology, multiscale biological annotations, and machine learning, this study provides a comprehensive framework for understanding the therapeutic effects of ECT in MDD. Our findings identify the SMN as a key, previously underappreciated substrate of ECT-related network reorganization. Rather than restoring baseline abnormalities toward a normative state, ECT was associated with a further reduction in SMN entropy, suggesting a treatment-related shift toward more constrained and functionally specialized sensorimotor integration. Importantly, this reorganization was selectively associated with improvements in suicidal ideation, psychic depression symptoms, and sleep disturbance, and was supported by convergent multiscale evidence consistent with serotonergic systems and inhibitory neuron-related transcriptional architecture. Furthermore, baseline edge-centric features demonstrated cross-site predictive utility for treatment response, underscoring their translational potential as prognostic biomarkers. Together, these findings extend current models of depression by emphasizing the role of sensorimotor network organization in the therapeutic effects of ECT and support edge-centric connectomics as a useful approach for characterizing treatment-related brain network reorganization.

## Data Availability

All data produced in the present study are available upon reasonable request to the authors

## Declarations

## Acknowledgements

This study was funded by the Shandong Provincial Natural Science Foundation (Kun Li, ZR2024QH385).

## Author contributions

Kexu Zhang: Conceptualization, Formal analysis, Methodology, Visualization, Writing - original draft, Writing - review & editing. Jiang Long: Methodology, Validation. Rui Li: Data curation, Validation. Xiaochi Yuan: Data curation, Software. Chenchen Zhang: Data curation. Ranran Xue: Data curation, Resources. Liju Qian: Data curation, Resources. Jiaojian Wang: Writing - review & editing. Wei Deng: Writing - review & editing. Yanghua Tian: Resources, Supervision. Kun Li: Conceptualization, Funding acquisition, Project administration, Writing - review & editing.

## Artificial Intelligence (AI) Statement

AI tools were used solely to refine, edit, and improve the language of human-written text. All scientific content, analyses, interpretations, and conclusions were developed and verified by the authors.

