## Supplementary Material for "Electroconvulsive Therapy Drives Sensorimotor Network Segregation in Depression: A Multiscale Edge-Centric Connectomic Study"

**Participants in the External Validation Cohort**

An independent validation cohort was obtained from Anhui Mental Health Center. Clinical diagnoses were determined by two trained psychiatrists based on the Structured Clinical Interview for DSM-IV (SCID-IV) in accordance with DSM-IV diagnostic criteria for major depressive disorder (MDD). Participants were excluded under the following conditions: (1) younger than 18 years or older than 65 years; (2) presence of substance abuse/dependence, severe medical conditions, neurological disorders, pregnancy, or comorbid psychiatric disorders such as schizophrenia, bipolar disorder, or personality disorders; (3) prior exposure to electroconvulsive therapy (ECT) within 6 months before enrollment; (4) contraindications to MRI examination or ECT treatment; (5) incomplete or excessive ECT treatment courses (<5 or >12 sessions).

**MRI Acquisition**

Neuroimaging data for the discovery cohort were acquired at the Department of Radiology, Shandong Daizhuang Hospital, using a 3.0-T Siemens MRI scanner. All participants underwent both high-resolution structural and resting-state functional MRI examinations. Structural images were obtained using a three-dimensional magnetization-prepared rapid gradient-echo (3D-MPRAGE) sequence with the following parameters: repetition time (TR) = 2500 ms, echo time (TE) = 2.25 ms, field of view (FOV) = 240 × 256 $mm^2$, acquisition matrix = 300 × 320, slice thickness = 0.8 mm, and 192 sagittal slices, resulting in an isotropic voxel resolution of 0.8 × 0.8 × 0.8 $mm^3$. Resting-state functional MRI (rs-fMRI) data were collected using a simultaneous multi-slice echo-planar imaging (SMS-EPI) sequence. Imaging parameters were as follows: TR = 1000 ms, TE = 33 ms, flip angle = 60°, FOV = 220 × 220 $mm^2$, matrix size = 90 × 90, slice thickness = 2.4 mm, voxel size = 2.44 × 2.44 × 2.4 $mm^3$, and 60 continuous axial slices. A multi-band acceleration factor of 6 was applied, and a total of 480 functional volumes were acquired for each participant.

MRI data for the independent validation cohort were acquired at Anhui Mental Health Center using a 3.0-T GE MRI scanner. High-resolution T1-weighted structural images were obtained using a sagittal three-dimensional inversion recovery fast spoiled gradient recalled (IR-FSPGR) sequence with the following parameters: repetition time (TR) = 8.16 ms, echo time (TE) = 3.28 ms, inversion time (TI) = 450 ms, flip angle = 12°, field of view (FOV) = 256 × 256 $mm^2$, acquisition matrix = 256 × 256, slice thickness = 1.0 mm, and 188 sagittal slices, yielding an isotropic voxel size of 1 × 1 × 1 $mm^3$. Resting-state functional images were acquired using a gradient-echo echo-planar imaging (EPI) sequence with the following parameters: repetition time (TR) = 2400 ms, echo time (TE) = 30 ms, flip angle = 90°, field of view (FOV) = 192 × 192 $mm^2$, matrix size = 64 × 64, slice thickness = 3.0 mm, voxel size = 3 × 3 × 3 $mm^3$, and 46 axial slices. A total of 217 functional volumes were collected for each participant.

**MRI Image Preprocessing**

All preprocessing steps for the resting-state functional MRI data were conducted utilizing the Data Processing & Analysis for Brain Imaging (DPABI) toolbox [1], which is built upon the Statistical Parametric Mapping (SPM12) software framework. To ensure magnetic field stabilization and allow participants to sufficiently acclimate to the scanning environment, the initial five temporal volumes of each functional run were discarded. The remaining images underwent slice-timing correction to compensate for intra-volume acquisition delays, followed by a realignment procedure to mitigate head motion artifacts. For precise spatial normalization, individual high-resolution T1-weighted structural images were first co-registered to the mean realigned functional volume. These co-registered structural images were subsequently segmented into distinct tissue probability maps (including white matter and cerebrospinal fluid) and spatially normalized to the standard Montreal Neurological Institute (MNI) template space. The non-linear deformation parameters derived from this structural-to-MNI normalization phase were then applied to warp the functional time series into the MNI space. Following spatial normalization, a multiple regression model was applied to regress out confounding nuisance covariates, including head motion parameters as well as the mean signals extracted from the white matter and cerebrospinal fluid (CSF). The residual functional images were then spatially smoothed utilizing a Gaussian kernel with a 6-mm full-width at half-maximum (FWHM) to enhance the signal-to-noise ratio.

Ultimately, a temporal band-pass filter (0.01–0.1 Hz) was applied to the time series to remove low-frequency scanner drift and high-frequency physiological interference.

**Computation of Edge-centric Normalized Entropy**

The theoretical framework and analytical pipeline for the edge-centric network analysis were adapted from previously established methodologies[2,3]. As outlined in the main text, regional blood-oxygen-level-dependent (BOLD) time series were unwrapped into dynamic co-fluctuation signals to construct the edge-centric functional connectivity (eFC) matrix. In general, eFC matrices are substantially larger than traditional node-centric functional connectivity matrices (246 nodes vs. 30258 edges). To alleviate the computational burden of clustering tens of thousands of edge observations, we implemented a dimensionality reduction procedure based on eigenvector decomposition. Specifically, we performed an eigen-decomposition of the individual eFC matrix and retained the top 50 eigenvectors. To ensure numerical stability before clustering, the coefficients of these eigenvectors were rescaled to the interval [-1, 1] by dividing each element by the maximum absolute magnitude within that vector.

The rescaled lower-dimensional edge features were subsequently partitioned into distinct, non-overlapping edge communities using a standard k-means clustering algorithm with Euclidean distance. We evaluated the optimal number of communities, k, utilizing the elbow method across a prespecified range (k = 2 to 20). Based on this analysis, k = 7 was identified as the optimal number of communities (Figure S6). During this phase, every edge connecting nodes $i$ and $j$ was assigned a deterministic community label $g_{ij} \in \{1,...,k\}$.

While edges are assigned to mutually exclusive clusters, the nodes themselves inherently exhibit an overlapping community structure, as each node participates in multiple edges. For a whole-brain network containing 246 regions (in our study, N = 246), each node $i$ connects to 245 edges (excluding self-connections). The participation proportion of region $i$ in a specific cluster $c$ was calculated as:

$$p_{ic} = \frac{1}{N-1}\sum_{j \neq i} \delta(g_{ij}, c)$$

where $\delta(x, y)$ denotes the Kronecker delta, which equals 1 if x = y and 0 otherwise. The vector $p_i = [p_{ii}, ..., p_{ik}]$ represents the community affiliation probability distribution for region $i$.

To quantitatively measure the extent of overlapping community affiliations for each region, we computed the Shannon entropy of its probability distribution. The raw community entropy $h_i$ for region $i$ is defined as:

$$h_i = -\sum_{c=1}^{k} p_{ic} \log_2 p_{ic}$$

To ensure the metric is bound to the interval [0, 1] and strictly comparable across different individuals, we divided the raw entropy by the theoretical maximum entropy:

$$H_i = \frac{h_i}{\log_2 k}$$

We refer to this normalized measure $H_i$ as the normalized entropy. Intuitively, if a region's edge affiliations are distributed uniformly across all functional networks, the normalized entropy approaches 1, indicating high integration and overlap. Conversely, if a region's edges are entirely confined to a single community, the normalized entropy approaches 0.

**Multivariate Predictive Modeling and Benchmark Validations**

To benchmark the prognostic utility of the edge-centric normalized entropy, two conventional regional and network topological metrics, fractional amplitude of low-frequency fluctuations (fALFF) and degree centrality (DC), were calculated using the DPABI toolbox.

First, fALFF was computed to measure the intensity of spontaneous regional brain activity. The preprocessed BOLD time series (strictly prior to temporal band-pass filtering) were transformed into the frequency domain utilizing a fast Fourier transform to obtain the power spectrum. The fALFF value for each region was calculated as the ratio of the power spectrum within the low-frequency band (0.01–0.08 Hz) to the total power across the entire detectable frequency range. Subsequently, DC was calculated to assess the functional hubness or global connectivity of each specific brain region. For each participant, a full-brain functional connectivity matrix was constructed by computing the Pearson correlation coefficients between the time series of all paired regions. To eliminate weak and potentially spurious connections, a predetermined correlation threshold ($r > 0.25$) was applied to the matrix, and the unweighted DC of a given region was defined as the sum of all suprathreshold connections associated with that node. Finally, both the region-level fALFF and DC values were normalized to z-scores for each individual to facilitate inter-subject comparability.

A rigorous predictive modeling framework was implemented using Support Vector Machine (SVM) and Support Vector Regression (SVR) algorithms. To unequivocally prevent data leakage and overfitting, feature selection was executed exclusively within the training set during each cross-validation fold or prior to the external validation phase. For the classification task (differentiating ECT responders from non-responders), candidate neuroimaging features were screened by comparing their baseline values between subgroups using independent two-sample *t*-tests or Mann-Whitney *U* tests, retaining only features exhibiting significant group differences ($p < 0.05$). These selected features, either independently or concatenated with clinical variables, were then fed into an SVM classifier with a linear kernel. The linear kernel was deliberately chosen to minimize the risk of overfitting inherent in high-dimensional neuroimaging datasets. The classification performance was systematically evaluated using accuracy, sensitivity, specificity, and the area under the receiver operating characteristic curve (AUC). Conversely, for the regression task (predicting the continuous HAMD reduction rate), feature screening was conducted using Pearson or Spearman correlation analyses between baseline features and the clinical reduction rate ($p < 0.05$). The retained features were utilized to train a linear-kernel SVR model, which maps the feature space to a continuous outcome by minimizing the generalized error bound. The regression performance was quantified via the correlation coefficient (r) between the predicted and actual HAMD reduction rates.

To rigorously ascertain the generalizability and robustness of these predictive models, two distinct validation strategies were adopted. Initially, an internal leave-one-out cross-validation (LOOCV) framework was applied within the discovery cohort (Site 1). In each iteration, data from one patient were withheld as the testing set, while the entire analytical pipeline—comprising feature selection and model training—was strictly confined to the remaining patients. Furthermore, to test the model's applicability across different clinical centers, an external independent validation scheme was implemented. Under this framework, the optimal features were selected and the final model was trained using the entirety of the Site 1 dataset. This trained model was then directly applied to the independent validation cohort (Site 2) without any further parameter tuning or feature re-selection.

**Cognitive Decoding via Neurosynth Meta-Analysis**

To robustly decode the functional and behavioral significance of the ECT-induced network reorganization, a spatial meta-analytic decoding approach was conducted utilizing the Neurosynth database. Specifically, the unthresholded region-level statistical map representing the longitudinal alterations in normalized entropy was spatially correlated with the meta-analytic term-based association maps available in the Neurosynth repository. This spatial correlation procedure generated a similarity coefficient (weight) for each term, quantifying the spatial overlap between our empirical entropy alteration pattern and the task-induced activation maps associated with diverse psychological domains. To ensure the neurobehavioral specificity of the decoding results, a stringent manual and programmatic filtering procedure was applied to the resulting list of terms. Specifically, anatomical descriptors (e.g., names of specific brain regions, sulci, or structural terms), non-specific methodological artifacts, and broad disease-related terms were systematically excluded to isolate purely cognitive and behavioral profiles. Following this curation process, the top 30 cognitive processes exhibiting the highest correlation weights were selectively extracted to phenomenologically characterize the macro-scale functional relevance of the observed network dynamics.

**Neurotransmitter Receptor and Transporter Mapping**

To investigate the neurochemical substrates underlying ECT-related network reorganization, spatial association analyses were performed between regional entropy alteration maps and whole-brain neurotransmitter receptor/transporter distributions derived from publicly available positron emission tomography (PET) atlases [4]. A total of 19 neurotransmitter receptor and transporter systems were included in the analysis, encompassing serotonin receptors and transporter (5-$HT_{1a}$, 5-$HT_{1b}$, 5-$HT_{2a}$, 5-$HT_4$, 5-$HT_6$, and 5-HTT), dopaminergic markers ($D_1$, $D_2$, DAT), noradrenergic transporter (NAT), histamine receptor ($H_3$), cholinergic markers ($\alpha_4\beta_2$, $M_1$, and VAChT), cannabinoid receptor type 1 ($CB_1$), μ-opioid receptor (MOR), N-methyl-D-aspartate receptor (NMDA), metabotropic glutamate receptor 5 ($mGluR_5$), and gamma-aminobutyric acid type A receptor ($GABA_A$).

All neurotransmitter maps were normalized to MNI standard space. To ensure spatial correspondence across modalities, both neurotransmitter density maps and entropy-derived spatial maps were parcellated according to the Brainnetome Atlas. Regional neurotransmitter densities were subsequently correlated with entropy alteration patterns across brain regions using Spearman correlation analysis. Multiple comparisons across neurotransmitter systems were controlled using false discovery rate (FDR) correction, with an adjusted threshold of FDR-corrected $p < 0.001$ established for statistical significance.

**Transcriptomic Association and Biological Enrichment Analyses**

To investigate the molecular architectures and genetic substrates underlying the macro-scale network reorganization, spatial transcriptomic association analyses were conducted using the Allen Human Brain Atlas (AHBA). The microarray gene expression data were preprocessed and parcellated to match the Brainnetome Atlas, yielding a region-by-gene expression matrix encompassing 15,633 genes across the whole brain [5]. Given the high dimensionality and spatial collinearity inherent in transcriptomic data, Partial Least Squares (PLS) regression was employed to identify robust linear combinations of gene expression patterns that covaried with the regional normalized entropy alterations. The first PLS component (PLS1), which accounted for the maximum covariance variance between the spatial gene expression profiles and the network dynamics, was selected for subsequent analyses. To ascertain the statistical significance of each gene's contribution to this primary transcriptomic signature, the PLS weights were strictly evaluated, and genes exhibiting a robust spatial association with an adjusted threshold of FDR-corrected $p<0.001$ were extracted. To further elucidate the functional significance of these ECT-related genes, comprehensive pathway enrichment analyses—incorporating Gene Ontology (GO) and Kyoto Encyclopedia of Genes and Genomes (KEGG) terms—were performed via the Metascape online platform (https://metascape.org/). Finally, to determine whether this identified gene set was disproportionately expressed in specific neural cell populations, a cell-type enrichment analysis was conducted utilizing reference transcriptomic profiles derived from single-cell transcriptomic studies summarized by Seidlitz et al. [6]. Both the pathway and cell-type enrichment results were rigorously constrained using a false discovery rate correction with a significance threshold of FDR-corrected $p < 0.001$.

**Associations Between Baseline Network Entropy and Clinical Symptoms**

To investigate the clinical relevance of baseline edge-centric community organization, partial correlation analyses were performed between normalized entropy values and symptom severity measures across all patients with MDD. Age, sex, and illness duration were included as covariates in all analyses.

At the network level, entropy within the visual network (VIS) was significantly negatively correlated with the Loss of Motivated Behavior subscale score (partial $r = -0.321$, $p = 0.004$), whereas entropy within the ventral attention network (VAN) showed a significant positive association with the same symptom dimension (partial r = 0.283, p = 0.012). In addition, entropy in the dorsal attention network (DAN) was associated with anxiety symptom severity, as reflected by positive correlations with the Anxiety subscale score (partial r = −0.250, p = 0.026). In contrast, entropy within the frontoparietal network (FPN) demonstrated significant positive correlations with both the Loss of Motivated Behavior subscale (partial r = 0.246, p = 0.029) and the Disturbed Thinking subscale (partial r = 0.282, p = 0.012). Furthermore, increased DMN entropy was positively associated with Anxiety subscale scores (partial r = 0.303, p = 0.007) and Hamilton Anxiety Rating Scale (HAMA) scores (partial r = 0.282, p = 0.012). Notably, entropy within the subcortical network (SCN) was positively correlated with suicidal ideation severity measured by the Columbia Suicide Severity Rating Scale (C-SSRS) (partial r = 0.222, p = 0.049), suggesting a potential involvement of subcortical network integration in suicidal symptomatology.

***References***


1 Yan C-G, Wang X-D, Zuo X-N, Zang Y-F. DPABI: Data processing & analysis for (resting-state) brain imaging. *Neuroinformatics* 2016; **14**: 339–351.

2 Faskowitz J, Esfahlani FZ, Jo Y, Sporns O, Betzel RF. Edge-centric functional network representations of human cerebral cortex reveal overlapping system-level architecture. *Nat Neurosci* 2020; **23**: 1644–1654.

3 Qin K, Ai C, Zhu P, Xiang J, Chen X, Zhang L *et al.* Edge-centric brain connectome representations reveal increased brain functional diversity of reward circuit in patients with major depressive disorder. *Biological Psychiatry* 2025; : S0006322325014301.

4 Dukart J, Holiga S, Rullmann M, Lanzenberger R, Hawkins PCT, Mehta MA *et al.* JuSpace: a tool for spatial correlation analyses of magnetic resonance imaging data with nuclear imaging derived neurotransmitter maps. *Hum Brain Mapp* 2020; **42**: 555–566.

5 Markello RD, Arnatkeviciute A, Poline J-B, Fulcher BD, Fornito A, Misic B. Standardizing workflows in imaging transcriptomics with the abagen toolbox. *Elife* 2021; **10**: e72129.

6 Seidlitz J, Nadig A, Liu S, Bethlehem RAI, Vértes PE, Morgan SE *et al.* Transcriptomic and cellular decoding of regional brain vulnerability to neurogenetic disorders. *Nat Commun* 2020; **11**: 3358.

Table S1. Demographic and clinical characteristics of pre-post comparison MDD patients in Site 1

| Characteristic | All patients | Responder | Non-responder |
|---|---|---|---|
| Number of subjects | 66 | 36 | 30 |
| Sex (Male/Female) | 24/42 | 12/24 | 12/18 |
| Age (years) | 21.49±7.71 | 22.75±8.97 | 19.97±5.65 |
| Duration of illness (months) | 46.56±36.26 | 43.94±40.16 | 49.70±31.33 |
| Number of ECT sessions | 8.00±3.51 | 8.44±3.17 | 7.47±3.88 |
| HAMD scores | | | |
| *Pre-ECT* | 34.49±6.80 | 33.03±6.10 | 36.23±7.28 |
| *Post-ECT* | 15.42 ±10.63 | 7.42±5.15 | 25.03±6.78 |
| *Statistics* | 16.10(T) | 24.63(T) | 9.34(T) |
| *p value* | <0.001 | <0.001 | <0.001 |
| HAMA scores | | | |
| *Pre-ECT* | 26.55±9.14 | 24.33±8.21 | 29.20±9.63 |
| *Post-ECT* | 13.08±9.74 | 6.39±4.87 | 21.20±7.86 |
| *Statistics* | 11.67(T) | 13.71(T) | 5.38(T) |
| *p value* | <0.001 | <0.001 | <0.001 |

*Note:* Data are presented as mean±standard deviation for continuous variables. Pre- versus post-treatment comparisons were performed using paired-sample t-tests, as the data satisfied the assumption of normality. T statistics are reported. HAMD, Hamilton Depression Rating Scale; HAMA, Hamilton Anxiety Rating Scale; ECT, electroconvulsive therapy.

Table S2 Brain regions exhibiting significant differences in normalized entropy between MDD patients and healthy controls.

| Node_id | Region | Network | MDD | HC | T value | p value | FDR-corrected p |
|---|---|---|---|---|---|---|---|
| 246 | Thalamus | SCN | 0.750±0.129 | 0.629±0.198 | 4.29 | <0.001 | <0.001 |
| 231 | Thalamus | SCN | 0.703±0.152 | 0.566±0.228 | 4.13 | <0.001 | <0.001 |
| 232 | Thalamus | SCN | 0.710±0.156 | 0.579±0.222 | 4.17 | <0.001 | <0.001 |
| 239 | Thalamus | SCN | 0.783±0.098 | 0.676±0.198 | 4.16 | <0.001 | <0.001 |
| 241 | Thalamus | SCN | 0.769±0.115 | 0.657±0.207 | 4.06 | <0.001 | <0.001 |
| 245 | Thalamus | SCN | 0.750±0.120 | 0.645±0.181 | 4.01 | <0.001 | <0.001 |
| 243 | Thalamus | SCN | 0.643±0.197 | 0.492±0.257 | 3.94 | <0.001 | <0.001 |
| 244 | Thalamus | SCN | 0.601±0.214 | 0.442±0.282 | 3.84 | <0.001 | <0.001 |
| 242 | Thalamus | SCN | 0.678±0.184 | 0.529±0.275 | 3.77 | <0.001 | <0.001 |
| 237 | Thalamus | SCN | 0.645±0.198 | 0.501±0.259 | 3.79 | <0.001 | <0.001 |
| 238 | Thalamus | SCN | 0.631±0.214 | 0.491±0.262 | 3.77 | <0.001 | <0.001 |
| 234 | Thalamus | SCN | 0.750±0.144 | 0.646±0.182 | 3.66 | <0.001 | <0.001 |
| 240 | Thalamus | SCN | 0.788±0.105 | 0.706±0.166 | 3.55 | <0.001 | 0.001 |
| 227 | Basal Ganglia | SCN | 0.611±0.212 | 0.473±0.256 | 3.55 | <0.001 | 0.001 |
| 228 | Basal Ganglia | SCN | 0.637±0.198 | 0.512±0.252 | 3.26 | 0.001 | 0.003 |
| 233 | Thalamus | SCN | 0.693±0.160 | 0.615±0.204 | 2.64 | 0.009 | 0.017 |
| 175 | Cingulate Gyrus | DMN | 0.626±0.129 | 0.526±0.174 | 3.61 | <0.001 | 0.019 |
| 223 | Basal Ganglia | SCN | 0.676±0.163 | 0.609±0.207 | 2.55 | 0.012 | 0.021 |

*Note:* Entropy values are presented as mean ± standard deviation. The Node_id corresponds to the regional index in the Brainnetome Atlas (246 regions). MDD, major depressive disorder; HC, healthy controls; SCN, subcortical network; DMN, default mode network; FDR, false discovery rate.

Table S3. Region-level longitudinal alterations of normalized entropy in all MDD patients following ECT.

| Node | Region | Network | Pre-ECT | Post-ECT | T value | p value | FDR-corrected p |
|---|---|---|---|---|---|---|---|
| 155 | Postcentral Gyrus | SMN | 0.716 ± 0.117 | 0.658 ± 0.149 | -2.945 | 0.004 | 0.101 |
| 218 | Hippocampus | SCN | 0.655 ± 0.176 | 0.727 ± 0.128 | 2.925 | 0.004 | 0.123 |
| 13 | Superior Frontal Gyrus | DMN | 0.695 ± 0.117 | 0.637 ± 0.129 | -2.833 | 0.005 | 0.247 |
| 159 | Postcentral Gyrus | SMN | 0.710 ± 0.116 | 0.658 ± 0.148 | -2.779 | 0.006 | 0.101 |
| 207 | lateral Occipital Cortex | VIS | 0.646 ± 0.162 | 0.577 ± 0.170 | -2.475 | 0.015 | 0.290 |
| 226 | Basal Ganglia | SCN | 0.709 ± 0.118 | 0.662 ± 0.154 | -2.379 | 0.019 | 0.212 |
| 176 | Cingulate Gyrus | DMN | 0.639 ± 0.131 | 0.583 ± 0.150 | -2.372 | 0.019 | 0.297 |
| 160 | Postcentral Gyrus | SMN | 0.705 ± 0.126 | 0.657 ± 0.139 | -2.366 | 0.020 | 0.172 |
| 30 | Inferior Frontal Gyrus | FPN | 0.660 ± 0.151 | 0.711 ± 0.137 | 2.320 | 0.022 | 0.230 |
| 65 | Paracentral Lobule | SMN | 0.739 ± 0.120 | 0.693 ± 0.147 | -2.288 | 0.024 | 0.172 |
| 229 | Basal Ganglia | SCN | 0.708 ± 0.122 | 0.656 ± 0.151 | -2.256 | 0.026 | 0.212 |
| 186 | Cingulate Gyrus | VAN | 0.769 ± 0.092 | 0.798 ± 0.083 | 2.246 | 0.026 | 0.661 |
| 156 | Postcentral Gyrus | SMN | 0.707 ± 0.123 | 0.664 ± 0.151 | -2.239 | 0.027 | 0.172 |
| 87 | Middle Temporal Gyrus | DMN | 0.655 ± 0.153 | 0.701 ± 0.115 | 2.226 | 0.028 | 0.297 |
| 195 | MedioVentral Occipital | VIS | 0.663 ± 0.164 | 0.601 ± 0.178 | -2.222 | 0.028 | 0.29 |
| 230 | Basal Ganglia | SCN | 0.718 ± 0.115 | 0.675 ± 0.136 | -2.220 | 0.028 | 0.212 |
| 114 | Parahippocampal Gyrus | VIS | 0.704 ± 0.142 | 0.752 ± 0.124 | 2.187 | 0.031 | 0.290 |
| 27 | Middle Frontal Gyrus | DMN | 0.662 ± 0.119 | 0.612 ± 0.148 | -2.186 | 0.031 | 0.297 |
| 34 | Inferior Frontal Gyrus | FPN | 0.644 ± 0.152 | 0.694 ± 0.129 | 2.181 | 0.031 | 0.230 |
| 14 | Superior Frontal Gyrus | DMN | 0.681 ± 0.103 | 0.640 ± 0.122 | -2.146 | 0.034 | 0.297 |
| 101 | Inferior Temporal Gyrus | LIM | 0.650 ± 0.175 | 0.706 ± 0.148 | 2.136 | 0.035 | 0.404 |
| 99 | Inferior Temporal Gyrus | FPN | 0.610 ± 0.138 | 0.656 ± 0.135 | 2.122 | 0.036 | 0.230 |
| 216 | Hippocampus | LIM | 0.690 ± 0.157 | 0.735 ± 0.142 | 2.122 | 0.036 | 0.404 |
| 49 | Orbital Gyrus | LIM | 0.756 ± 0.107 | 0.722 ± 0.118 | -2.111 | 0.037 | 0.404 |
| 67 | Paracentral Lobule | SMN | 0.734 ± 0.117 | 0.686 ± 0.154 | -2.109 | 0.037 | 0.196 |
| 130 | Superior Parietal Lobule | DAN | 0.680 ± 0.137 | 0.633 ± 0.146 | -2.107 | 0.037 | 0.720 |
| 198 | MedioVentral Occipital | VIS | 0.688 ± 0.145 | 0.636 ± 0.152 | -2.089 | 0.039 | 0.29 |
| 242 | Thalamus | SCN | 0.683 ± 0.189 | 0.736 ± 0.139 | 2.080 | 0.040 | 0.237 |
| 83 | Middle Temporal Gyrus | DMN | 0.613 ± 0.161 | 0.661 ± 0.130 | 2.055 | 0.042 | 0.297 |
| 142 | Inferior Parietal Lobule | FPN | 0.580 ± 0.178 | 0.643 ± 0.184 | 2.052 | 0.042 | 0.230 |
| 157 | Postcentral Gyrus | SMN | 0.706 ± 0.113 | 0.666 ± 0.146 | -2.046 | 0.043 | 0.196 |
| 52 | Orbital Gyrus | DMN | 0.686 ± 0.128 | 0.728 ± 0.115 | 2.023 | 0.045 | 0.297 |

*Note:* Entropy values are presented as mean ± standard deviation. The Node index corresponds to the specific region in the Brainnetome Atlas (246 ROIs). MDD, major depressive disorder; ECT, electroconvulsive therapy; FDR, false discovery rate; SMN, somatomotor network; SCN, subcortical network; DMN, default mode network; VIS, visual network; FPN, frontoparietal network; VAN, ventral attention network; LIM, limbic network; DAN, dorsal attention network.

Table S4. Region-level longitudinal alterations of normalized entropy in responders following ECT.

| Node | Region | Network | Pre-ECT | Post-ECT | T value | p value | FDR-corrected p |
|---|---|---|---|---|---|---|---|
| 159 | Postcentral Gyrus | SMN | 0.720 ± 0.135 | 0.632 ± 0.163 | -3.423 | 0.001 | 0.034 |
| 142 | Inferior Parietal Lobule | FPN | 0.538 ± 0.205 | 0.669 ± 0.157 | 3.268 | 0.002 | 0.038 |
| 160 | Postcentral Gyrus | SMN | 0.730 ± 0.121 | 0.651 ± 0.148 | -2.942 | 0.004 | 0.067 |
| 87 | Middle Temporal Gyrus | DMN | 0.645 ± 0.146 | 0.715 ± 0.090 | 2.700 | 0.009 | 0.293 |
| 30 | Inferior Frontal Gyrus | FPN | 0.667 ± 0.147 | 0.745 ± 0.102 | 2.693 | 0.009 | 0.099 |
| 195 | MedioVentral Occipital | VIS | 0.684 ± 0.162 | 0.583 ± 0.181 | -2.661 | 0.010 | 0.206 |
| 130 | Superior Parietal Lobule | DAN | 0.691 ± 0.123 | 0.617 ± 0.135 | -2.636 | 0.010 | 0.292 |
| 72 | Superior Temporal Gyrus | SMN | 0.712 ± 0.083 | 0.650 ± 0.145 | -2.623 | 0.011 | 0.067 |
| 155 | Postcentral Gyrus | SMN | 0.734 ± 0.100 | 0.659 ± 0.155 | -2.621 | 0.011 | 0.067 |
| 164 | Insular Gyrus | SMN | 0.735 ± 0.099 | 0.666 ± 0.169 | -2.590 | 0.012 | 0.067 |
| 163 | Insular Gyrus | SMN | 0.741 ± 0.062 | 0.678 ± 0.152 | -2.564 | 0.013 | 0.067 |
| 208 | lateral Occipital Cortex | VIS | 0.641 ± 0.167 | 0.548 ± 0.171 | -2.532 | 0.014 | 0.206 |
| 34 | Inferior Frontal Gyrus | FPN | 0.614 ± 0.170 | 0.692 ± 0.139 | 2.504 | 0.015 | 0.108 |
| 141 | Inferior Parietal Lobule | DMN | 0.577 ± 0.165 | 0.654 ± 0.141 | 2.433 | 0.018 | 0.293 |
| 73 | Superior Temporal Gyrus | SMN | 0.726 ± 0.079 | 0.667 ± 0.154 | -2.423 | 0.018 | 0.073 |
| 162 | Postcentral Gyrus | SMN | 0.685 ± 0.126 | 0.607 ± 0.157 | -2.420 | 0.018 | 0.073 |
| 122 | posterior Superior Temporal | DMN | 0.657 ± 0.176 | 0.737 ± 0.111 | 2.403 | 0.019 | 0.293 |
| 225 | Basal Ganglia | SCN | 0.713 ± 0.107 | 0.654 ± 0.128 | -2.369 | 0.021 | 0.475 |
| 55 | Precentral Gyrus | DAN | 0.770 ± 0.086 | 0.725 ± 0.103 | -2.259 | 0.027 | 0.293 |
| 192 | MedioVentral Occipital | VIS | 0.668 ± 0.193 | 0.570 ± 0.206 | -2.253 | 0.028 | 0.218 |
| 7 | Superior Frontal Gyrus | DAN | 0.742 ± 0.096 | 0.684 ± 0.134 | -2.199 | 0.031 | 0.293 |
| 117 | Parahippocampal Gyrus | LIM | 0.452 ± 0.253 | 0.574 ± 0.247 | 2.196 | 0.032 | 0.992 |
| 18 | Middle Frontal Gyrus | FPN | 0.679 ± 0.140 | 0.742 ± 0.119 | 2.164 | 0.034 | 0.187 |
| 196 | MedioVentral Occipital | VIS | 0.662 ± 0.186 | 0.576 ± 0.183 | -2.143 | 0.036 | 0.218 |
| 207 | lateral Occipital Cortex | VIS | 0.630 ± 0.181 | 0.545 ± 0.184 | -2.137 | 0.036 | 0.218 |
| 226 | Basal Ganglia | SCN | 0.709 ± 0.131 | 0.655 ± 0.150 | -2.127 | 0.037 | 0.475 |
| 54 | Precentral Gyrus | SMN | 0.719 ± 0.107 | 0.651 ± 0.187 | -2.124 | 0.037 | 0.111 |
| 176 | Cingulate Gyrus | DMN | 0.654 ± 0.114 | 0.592 ± 0.148 | -2.118 | 0.038 | 0.436 |
| 71 | Superior Temporal Gyrus | SMN | 0.731 ± 0.079 | 0.679 ± 0.145 | -2.089 | 0.041 | 0.111 |
| 61 | Precentral Gyrus | VAN | 0.672 ± 0.089 | 0.629 ± 0.136 | -2.074 | 0.042 | 0.35 |
| 9 | Superior Frontal Gyrus | SMN | 0.755 ± 0.083 | 0.704 ± 0.133 | -2.072 | 0.042 | 0.111 |
| 65 | Paracentral Lobule | SMN | 0.744 ± 0.114 | 0.691 ± 0.157 | -2.049 | 0.044 | 0.111 |
| 53 | Precentral Gyrus | SMN | 0.748 ± 0.101 | 0.691 ± 0.158 | -2.043 | 0.045 | 0.111 |
| 184 | Cingulate Gyrus | VAN | 0.743 ± 0.069 | 0.697 ± 0.133 | -2.026 | 0.047 | 0.350 |
| 173 | Insular Gyrus | VAN | 0.644 ± 0.116 | 0.592 ± 0.150 | -2.024 | 0.047 | 0.350 |
| 59 | Precentral Gyrus | SMN | 0.679 ± 0.180 | 0.595 ± 0.191 | -2.011 | 0.048 | 0.111 |

*Note:* Entropy values are presented as mean ± standard deviation. The Node index corresponds to the specific region in the Brainnetome Atlas (246 ROIs). ECT, electroconvulsive therapy; FDR, false discovery rate; SMN, somatomotor network; SCN, subcortical network; DMN, default mode network; VIS, visual network; FPN, frontoparietal network; VAN, ventral attention network; LIM, limbic network; DAN, dorsal attention network.

Table S5. Region-level longitudinal alterations of normalized entropy in non-responders following ECT.

| Node | Region | Network | Pre-ECT | Post-ECT | T value | p value | FDR-corrected p |
|---|---|---|---|---|---|---|---|
| 120 | Parahippocampal Gyrus | VIS | 0.708 ± 0.123 | 0.773 ± 0.090 | 2.839 | 0.006 | 0.191 |
| 27 | Middle Frontal Gyrus | DMN | 0.687 ± 0.106 | 0.607 ± 0.132 | -2.668 | 0.010 | 0.354 |
| 13 | Superior Frontal Gyrus | DMN | 0.700 ± 0.141 | 0.631 ± 0.106 | -2.345 | 0.023 | 0.354 |
| 101 | Inferior Temporal Gyrus | LIM | 0.617 ± 0.203 | 0.716 ± 0.166 | 2.268 | 0.027 | 0.434 |
| 218 | Hippocampus | SCN | 0.636 ± 0.217 | 0.724 ± 0.104 | 2.186 | 0.033 | 0.742 |
| 121 | posterior Superior Temporal Sulcus | DMN | 0.727 ± 0.137 | 0.658 ± 0.134 | -2.133 | 0.038 | 0.354 |
| 165 | Insular Gyrus | DMN | 0.665 ± 0.202 | 0.746 ± 0.113 | 2.084 | 0.042 | 0.354 |
| 112 | Parahippocampal Gyrus | LIM | 0.681 ± 0.193 | 0.759 ± 0.095 | 2.067 | 0.044 | 0.434 |
| 74 | Superior Temporal Gyrus | SMN | 0.701 ± 0.126 | 0.745 ± 0.081 | 2.044 | 0.046 | 0.738 |
| 163 | Insular Gyrus | SMN | 0.719 ± 0.110 | 0.761 ± 0.079 | 2.041 | 0.046 | 0.738 |
| 102 | Inferior Temporal Gyrus | LIM | 0.630 ± 0.201 | 0.716 ± 0.148 | 2.037 | 0.047 | 0.434 |

*Note:* Entropy values are presented as mean ± standard deviation. The Node index corresponds to the specific region in the Brainnetome Atlas (246 ROIs). ECT, electroconvulsive therapy; FDR, false discovery rate; SMN, somatomotor network; SCN, subcortical network; DMN, default mode network; VIS, visual network; LIM, limbic network.

Table S6. Region-level longitudinal alterations of normalized entropy following ECT in the independent validation dataset.

| Node | Region | Network | Pre-ECT | Post-ECT | T value | p value | FDR-corrected p |
|---|---|---|---|---|---|---|---|
| 245 | Thalamus | SCN | 0.749 ± 0.102 | 0.637 ± 0.127 | -4.141 | <0.001 | 0.004 |
| 71 | Superior Temporal Gyrus | SMN | 0.722 ± 0.098 | 0.638 ± 0.100 | -4.041 | <0.001 | 0.005 |
| 218 | Hippocampus | SCN | 0.636 ± 0.161 | 0.466 ± 0.195 | -3.902 | <0.001 | 0.005 |
| 211 | Amygdala | SCN | 0.666 ± 0.149 | 0.551 ± 0.220 | -3.406 | 0.001 | 0.013 |
| 216 | Hippocampus | SCN | 0.693 ± 0.135 | 0.578 ± 0.148 | -3.358 | 0.001 | 0.013 |
| 172 | Insular Gyrus | SMN | 0.698 ± 0.145 | 0.613 ± 0.147 | -3.054 | 0.004 | 0.056 |
| 243 | Thalamus | SCN | 0.677 ± 0.148 | 0.545 ± 0.198 | -2.983 | 0.004 | 0.031 |
| 217 | Hippocampus | SCN | 0.627 ± 0.168 | 0.499 ± 0.227 | -2.884 | 0.006 | 0.034 |
| 117 | Parahippocampal Gyrus | LIM | 0.677 ± 0.153 | 0.572 ± 0.199 | -2.852 | 0.006 | 0.112 |
| 109 | Parahippocampal Gyrus | LIM | 0.626 ± 0.131 | 0.509 ± 0.206 | -2.741 | 0.008 | 0.112 |
| 246 | Thalamus | SCN | 0.730 ± 0.120 | 0.655 ± 0.130 | -2.731 | 0.009 | 0.044 |
| 183 | Cingulate Gyrus | VAN | 0.754 ± 0.107 | 0.682 ± 0.121 | -2.704 | 0.009 | 0.094 |
| 80 | Superior Temporal Gyrus | SMN | 0.730 ± 0.160 | 0.639 ± 0.165 | -2.653 | 0.010 | 0.098 |
| 114 | Parahippocampal Gyrus | VIS | 0.655 ± 0.196 | 0.532 ± 0.215 | -2.652 | 0.010 | 0.305 |
| 174 | Insular Gyrus | VAN | 0.696 ± 0.129 | 0.623 ± 0.137 | -2.624 | 0.011 | 0.094 |
| 232 | Thalamus | SCN | 0.708 ± 0.113 | 0.623 ± 0.161 | -2.594 | 0.012 | 0.048 |
| 73 | Superior Temporal Gyrus | SMN | 0.710 ± 0.122 | 0.651 ± 0.094 | -2.591 | 0.012 | 0.098 |
| 215 | Hippocampus | SCN | 0.682 ± 0.173 | 0.572 ± 0.177 | -2.562 | 0.013 | 0.048 |
| 238 | Thalamus | SCN | 0.672 ± 0.134 | 0.568 ± 0.187 | -2.557 | 0.013 | 0.048 |
| 110 | Parahippocampal Gyrus | LIM | 0.636 ± 0.139 | 0.541 ± 0.176 | -2.555 | 0.013 | 0.121 |
| 241 | Thalamus | SCN | 0.694 ± 0.128 | 0.606 ± 0.151 | -2.483 | 0.016 | 0.053 |
| 171 | Insular Gyrus | VAN | 0.691 ± 0.130 | 0.628 ± 0.110 | -2.463 | 0.017 | 0.094 |
| 78 | Superior Temporal Gyrus | LIM | 0.688 ± 0.158 | 0.597 ± 0.186 | -2.413 | 0.019 | 0.130 |
| 208 | lateral Occipital Cortex | VIS | 0.700 ± 0.138 | 0.611 ± 0.177 | -2.391 | 0.020 | 0.305 |
| 212 | Amygdala | SCN | 0.697 ± 0.145 | 0.610 ± 0.181 | -2.388 | 0.020 | 0.061 |
| 184 | Cingulate Gyrus | VAN | 0.746 ± 0.096 | 0.688 ± 0.113 | -2.376 | 0.021 | 0.094 |
| 145 | Inferior Parietal Lobule | VAN | 0.705 ± 0.125 | 0.653 ± 0.105 | -2.372 | 0.021 | 0.094 |
| 168 | Insular Gyrus | VAN | 0.716 ± 0.113 | 0.659 ± 0.126 | -2.347 | 0.023 | 0.094 |
| 237 | Thalamus | SCN | 0.670 ± 0.140 | 0.582 ± 0.166 | -2.316 | 0.024 | 0.068 |
| 10 | Superior Frontal Gyrus | SMN | 0.728 ± 0.135 | 0.666 ± 0.104 | -2.304 | 0.025 | 0.128 |
| 163 | Insular Gyrus | SMN | 0.691 ± 0.113 | 0.637 ± 0.116 | -2.303 | 0.025 | 0.128 |
| 180 | Cingulate Gyrus | VAN | 0.749 ± 0.113 | 0.678 ± 0.152 | -2.284 | 0.026 | 0.094 |
| 214 | Amygdala | SCN | 0.608 ± 0.179 | 0.511 ± 0.188 | -2.272 | 0.027 | 0.070 |
| 244 | Thalamus | SCN | 0.622 ± 0.167 | 0.528 ± 0.218 | -2.202 | 0.032 | 0.075 |
| 242 | Thalamus | SCN | 0.611 ± 0.194 | 0.503 ± 0.246 | -2.184 | 0.033 | 0.075 |
| 70 | Superior Temporal Gyrus | LIM | 0.703 ± 0.154 | 0.631 ± 0.176 | -2.142 | 0.037 | 0.198 |
| 76 | Superior Temporal Gyrus | SMN | 0.697 ± 0.172 | 0.631 ± 0.131 | -2.141 | 0.037 | 0.128 |
| 157 | Postcentral Gyrus | SMN | 0.708 ± 0.121 | 0.648 ± 0.108 | -2.139 | 0.037 | 0.128 |
| 120 | Parahippocampal Gyrus | VIS | 0.639 ± 0.192 | 0.550 ± 0.199 | -2.103 | 0.040 | 0.401 |
| 35 | Inferior Frontal Gyrus | DMN | 0.651 ± 0.134 | 0.715 ± 0.108 | 2.102 | 0.040 | 0.851 |
| 74 | Superior Temporal Gyrus | SMN | 0.718 ± 0.122 | 0.669 ± 0.097 | -2.082 | 0.042 | 0.128 |
| 9 | Superior Frontal Gyrus | SMN | 0.721 ± 0.130 | 0.657 ± 0.132 | -2.069 | 0.043 | 0.128 |
| 57 | Precentral Gyrus | SMN | 0.702 ± 0.152 | 0.634 ± 0.139 | -2.063 | 0.044 | 0.128 |
| 240 | Thalamus | SCN | 0.649 ± 0.180 | 0.552 ± 0.198 | -2.061 | 0.044 | 0.094 |
| 167 | Insular Gyrus | VAN | 0.722 ± 0.125 | 0.661 ± 0.152 | -2.01 | 0.049 | 0.154 |

*Note:* Entropy values are presented as mean ± standard deviation. The Node index corresponds to the specific region in the Brainnetome Atlas (246 ROIs). ECT, electroconvulsive therapy; FDR, false discovery rate; SCN, subcortical network; SMN, somatomotor network; LIM, limbic network; VAN, ventral attention network; VIS, visual network; DMN, default mode network.

Table S7. Classification performance of different feature combinations for predicting ECT efficacy.

| Model | LOOCV | | | | External | | | |
|---|---|---|---|---|---|---|---|---|
| | AUC | ACC | Sensitivity | Specificity | AUC | ACC | Sensitivity | Specificity |
| Clinical Only | 0.605 | 0.591 | 0.611 | 0.567 | 0.5901 | 0.533 | 0.565 | 0.429 |
| Clinical + Entropy | **0.759** | **0.727** | **0.750** | **0.700** | **0.5963** | **0.633** | **0.652** | **0.571** |
| Clinical + fALFF | 0.740 | 0.606 | 0.583 | 0.633 | 0.5031 | 0.433 | 0.435 | 0.429 |
| Clinical + DC | 0.744 | 0.591 | 0.639 | 0.533 | 0.5652 | 0.600 | 0.652 | 0.429 |

*Note:* LOOCV, leave-one-out cross-validation; AUC, area under the curve; ACC, accuracy; fALFF, fractional amplitude of low-frequency fluctuation; DC, degree centrality.

Table S8. Top 30 cognitive terms associated with the spatial pattern of ECT-induced entropy alterations.

| Rank | Cognitive Concept | Weight | Rank | Cognitive Concept | Weight |
|---|---|---|---|---|---|
| 1 | sensorimotor | -0.3543 | 16 | semantic | 0.2217 |
| 2 | somatosensory | -0.3439 | 17 | recognizing | 0.2187 |
| 3 | recognition | 0.3094 | 18 | face recognition | 0.2184 |
| 4 | movement | -0.2991 | 19 | word form | 0.2182 |
| 5 | extension | -0.2905 | 20 | concepts | 0.2172 |
| 6 | face | 0.2756 | 21 | matching task | 0.2165 |
| 7 | memory | 0.2653 | 22 | knowledge | 0.2154 |
| 8 | encoding | 0.2618 | 23 | conceptual | 0.2144 |
| 9 | identity | 0.2585 | 24 | memory encoding | 0.2129 |
| 10 | unfamiliar | 0.2526 | 25 | word | 0.2122 |
| 11 | term memory | 0.2469 | 26 | visual word | 0.2117 |
| 12 | coordination | -0.2446 | 27 | subsequent memory | 0.2081 |
| 13 | recognize | 0.2437 | 28 | familiarity | 0.2053 |
| 14 | familiar | 0.2413 | 29 | long term | 0.2048 |
| 15 | retrieval | 0.2273 | 30 | binding | 0.2043 |

Table S9. Spatial correlations between ECT-induced entropy alterations and the cortical distributions of neurotransmitter receptors and transporters.

| Neurotransmitter | ρ | p | FDR-corrected p |
|---|---|---|---|
| $5HT_{1a}$ | 0.285 | <0.001 | <0.001 |
| $5HT_{1b}$ | -0.215 | 0.001 | 0.002 |
| $5HT_{2a}$ | 0.080 | 0.214 | 0.312 |
| $5HT_4$ | 0.307 | <0.001 | <0.001 |
| $5HT_6$ | -0.085 | 0.182 | 0.306 |
| 5HTT | -0.083 | 0.193 | 0.306 |
| $D_1$ | -0.009 | 0.889 | 0.892 |
| $D_2$ | 0.224 | <0.001 | 0.002 |
| DAT | 0.069 | 0.281 | 0.355 |
| NAT | -0.264 | <0.001 | <0.001 |
| $H_3$ | -0.231 | <0.001 | 0.001 |
| $\alpha_4\beta_2$ | -0.111 | 0.083 | 0.175 |
| $M_1$ | -0.018 | 0.777 | 0.868 |
| VAChT | -0.161 | 0.011 | 0.027 |
| $CB_1$ | 0.102 | 0.110 | 0.210 |
| MOR | 0.176 | 0.006 | 0.015 |
| NMDA | -0.073 | 0.251 | 0.340 |
| $mGluR_5$ | -0.055 | 0.387 | 0.459 |
| GABAA | 0.009 | 0.892 | 0.892 |

Table S10 Gene Ontology and KEGG pathway enrichment results of PLS1 significant genes

| GO | Category | Description | Count | Ratio(%) | Log10(q) |
|---|---|---|---|---|---|
| GO:0050804 | Biological Process | Modulation of chemical synaptic transmission | 38 | 6.31 | -6.57 |
| GO:0098793 | Cellular Component | Presynapse | 41 | 6.81 | -5.92 |
| GO:0030425 | Cellular Component | Dendrite | 39 | 6.48 | -5.92 |
| GO:0099536 | Biological Process | Synaptic signaling | 33 | 5.48 | -5.35 |
| GO:0019904 | Molecular Function | Protein domain specific binding | 38 | 6.31 | -5.09 |
| hsa04360 | KEGG Pathway | Axon guidance | 19 | 3.16 | -5 |
| GO:0034702 | Cellular Component | Monoatomic ion channel complex | 27 | 4.49 | -4.89 |
| hsa05200 | KEGG Pathway | Pathways in cancer | 32 | 5.32 | -4.26 |
| GO:0050808 | Biological Process | Synapse organization | 25 | 4.15 | -3.6 |
| GO:0141124 | Biological Process | Intracellular signaling cassette | 39 | 6.48 | -3.59 |
| GO:0030900 | Biological Process | Forebrain development | 26 | 4.32 | -3.53 |
| GO:0034762 | Biological Process | Regulation of transmembrane transport | 27 | 4.49 | -3.44 |
| GO:0003012 | Biological Process | Muscle system process | 23 | 3.82 | -3.4 |
| GO:0031175 | Biological Process | Neuron projection development | 36 | 5.98 | -3.38 |
| GO:0060341 | Biological Process | Regulation of cellular localization | 30 | 4.98 | -3.23 |

*Note:* The statistical significance is represented by Log10(q), where the q-value is the false discovery rate (FDR)-adjusted p-value. GO, Gene Ontology; KEGG, Kyoto Encyclopedia of Genes and Genomes.

Table S11 Overlap between the PLS1 significant genes and canonical brain cell-type markers.

| Cell types | Number of common genes | p | FDR-corrected p |
|---|---|---|---|
| Astrocytes | 38 | 0.318 | 0.557 |
| Endothelial cells | 28 | 0.966 | 0.966 |
| Microglia | 26 | 0.894 | 0.966 |
| Excitatory neurons | 55 | 0.002 | 0.007 |
| Inhibitory neurons | 57 | <0.001 | <0.001 |
| Oligodendrocyte precursor cells | 11 | 0.018 | 0.042 |
| Oligodendrocytes | 30 | 0.877 | 0.966 |

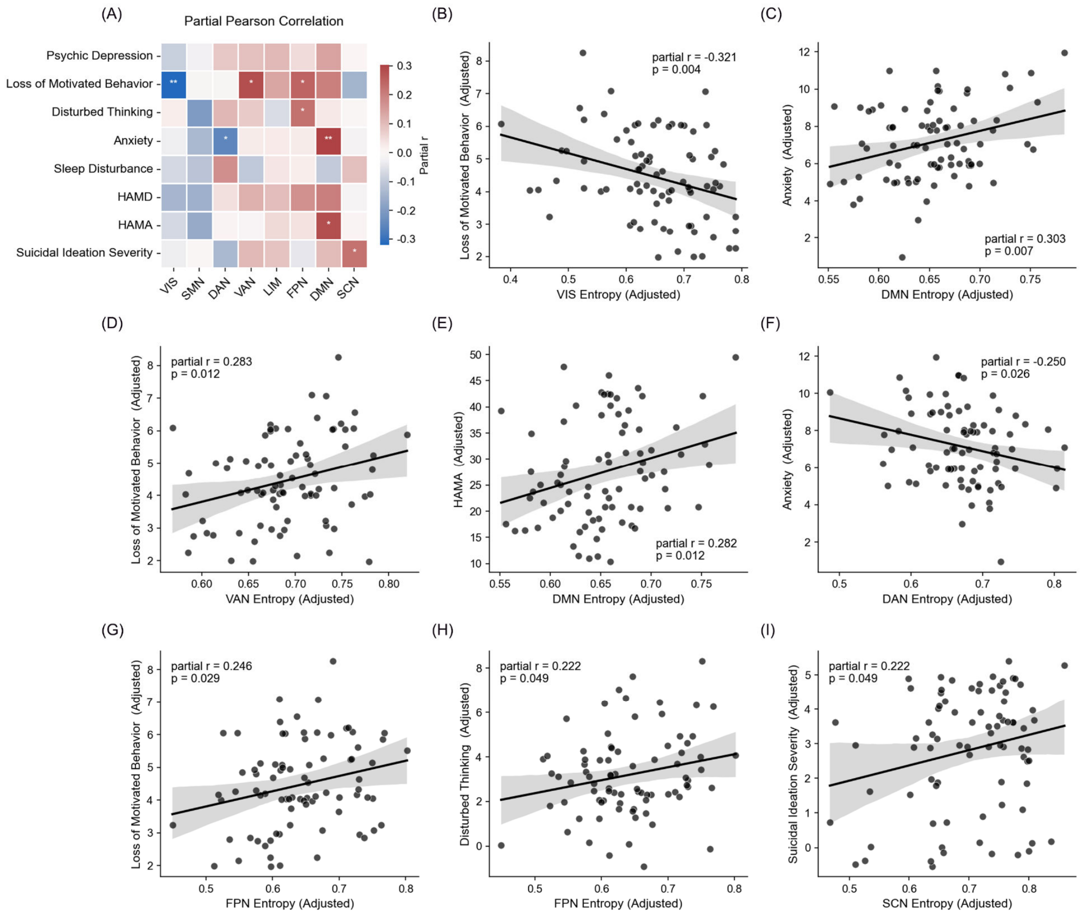


Figure S1. Baseline correlations between network-level normalized entropy and clinical symptoms in MDD patients. (A) Heatmap showing partial Pearson correlation coefficients between baseline (Pre-ECT) normalized entropy across eight functional networks and clinical symptom scores. Significant correlations are marked with asterisks (*$p < 0.05$, **$p < 0.01$). (B–I) Scatter plots illustrating the corresponding significant partial correlations identified in the heatmap. Entropy and symptom scores are adjusted for covariates. MDD, major depressive disorder; ECT, electroconvulsive therapy; HAMD, Hamilton Depression Rating Scale; HAMA, Hamilton Anxiety Rating Scale.

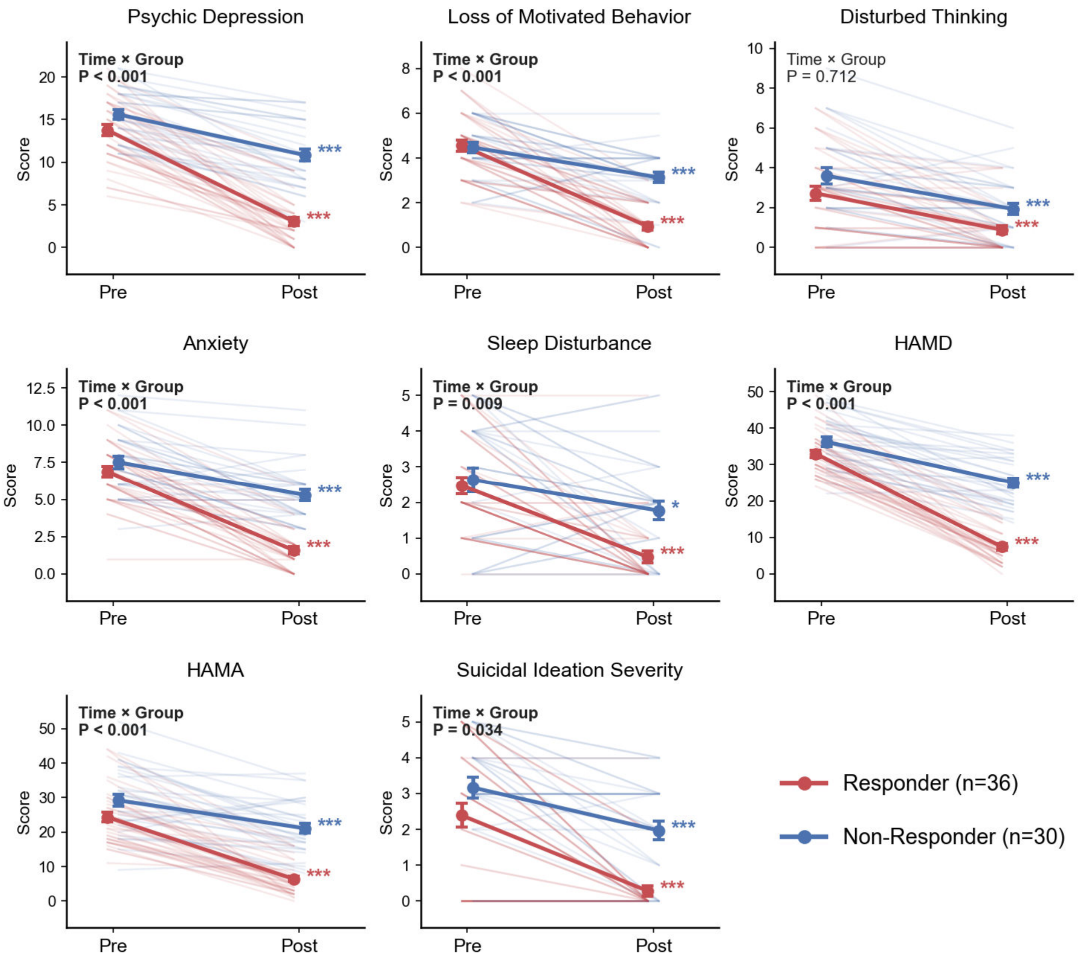


Figure S2. Longitudinal improvements in clinical symptoms following ECT in MDD patients. Line plots illustrate the changes in diverse clinical scores from pre-ECT to post-ECT for the Responder (red, n=36) and Non-Responder (blue, n=30) subgroups. MDD, major depressive disorder; ECT, electroconvulsive therapy; HAMD, Hamilton Depression Rating Scale; HAMA, Hamilton Anxiety Rating Scale.

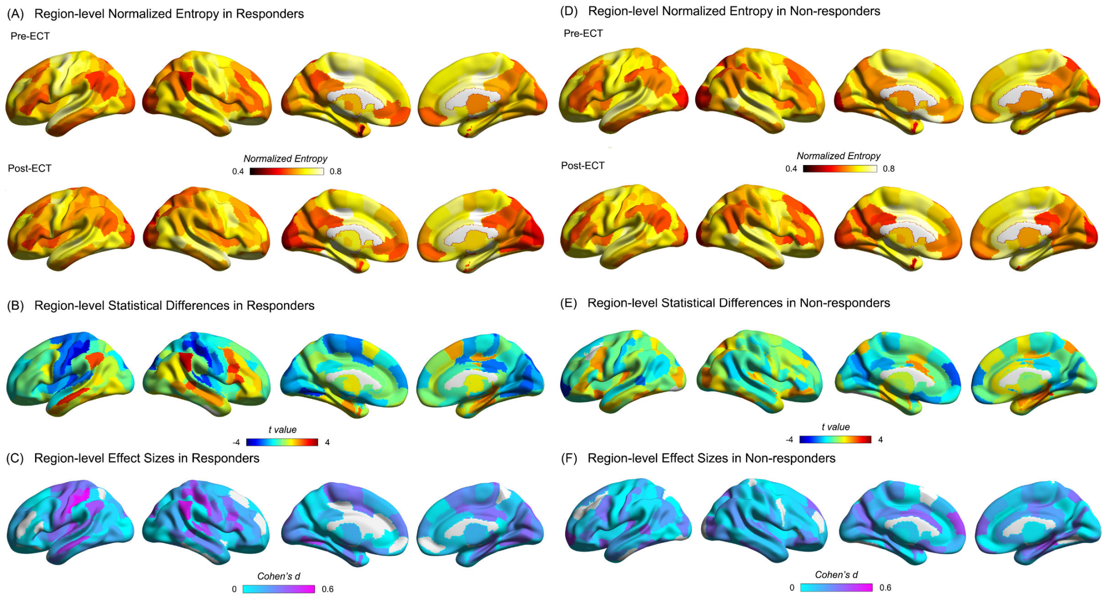


Figure S3. Region-level longitudinal alterations in edge-centric network entropy within the Responder and Non-responder subgroups. (A–C) Brain surface maps for the Responder subgroup showing: (A) the spatial distribution of normalized entropy before (Pre-ECT) and after (Post-ECT) treatment; (B) the unthresholded statistical differences (t-values) of longitudinal entropy changes; and (C) the corresponding effect sizes (Cohen's d). (D–F) Brain surface maps for the Non-responder subgroup showing: (D) the spatial distribution of normalized entropy pre- and post-ECT; (E) the unthresholded statistical differences (t-values); and (F) the corresponding effect sizes (Cohen's d). ECT, electroconvulsive therapy.

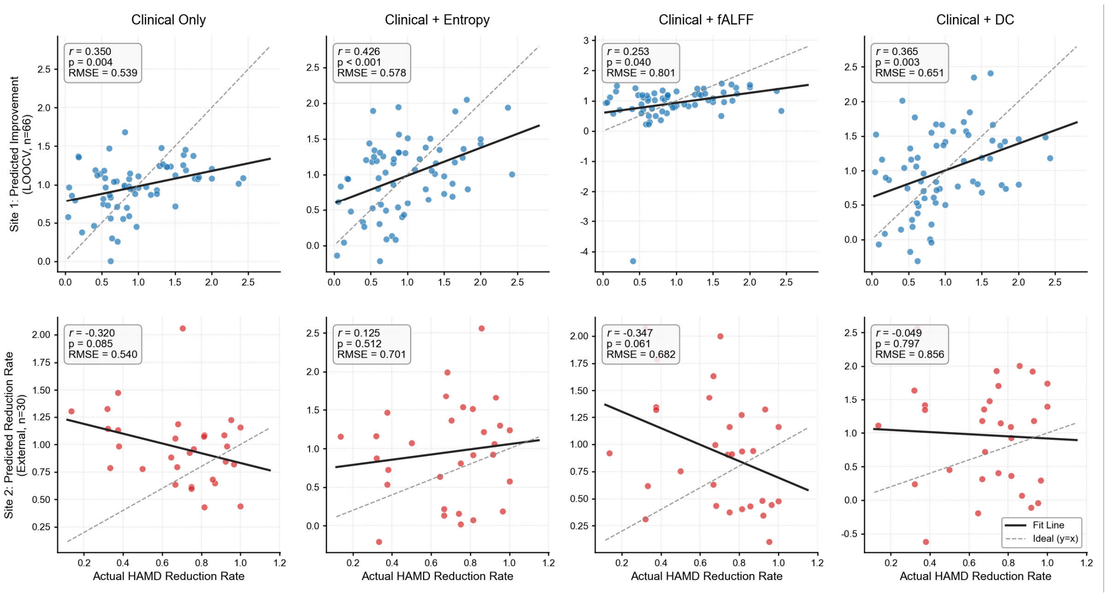


Figure S4. Predictive performance of diverse feature combinations for estimating depressive symptom reduction rates. Scatter plots illustrate the correlations between the actual (observed) and predicted HAMD reduction rates following ECT. The top row displays the internal leave-one-out cross-validation (LOOCV) results within the discovery cohort (Site 1, n=66). The bottom row presents the external validation results (Site 2, n=30) using regression models trained on Site 1. Four feature combinations were evaluated: Clinical Only, Clinical + Entropy, Clinical + fALFF, and Clinical + DC. ECT, electroconvulsive therapy; HAMD, Hamilton Depression Rating Scale; LOOCV, leave-one-out cross-validation; RMSE, root mean square error; fALFF, fractional amplitude of low-frequency fluctuation; DC, degree centrality.

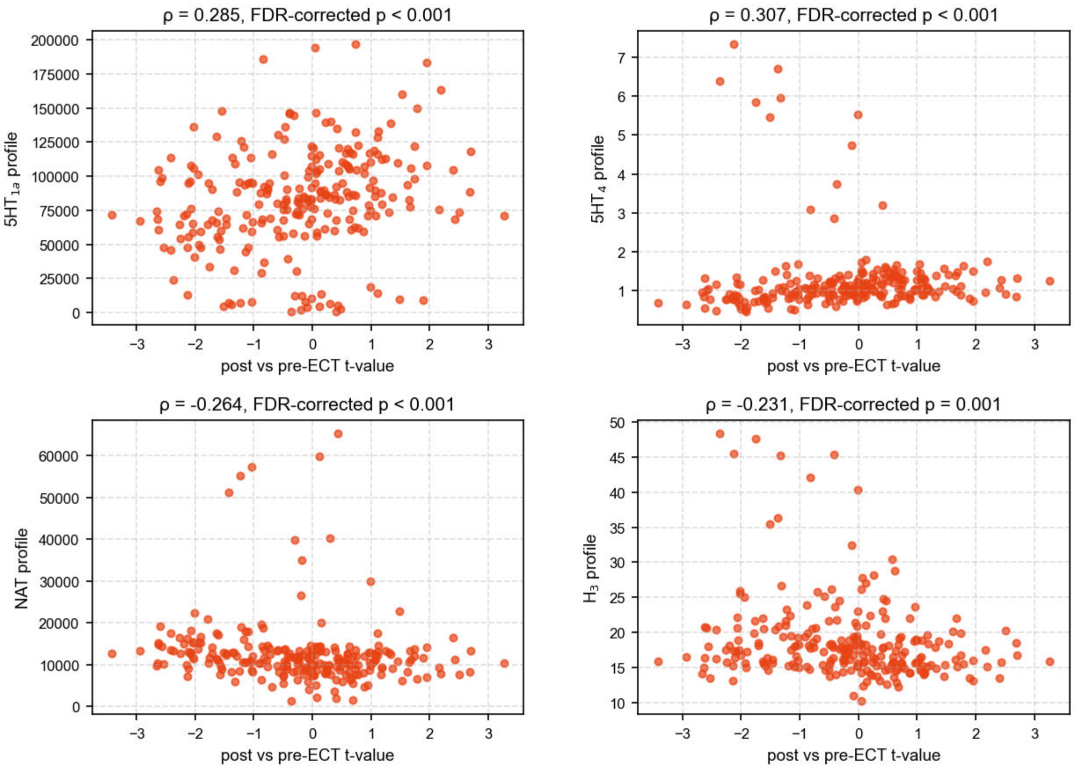


Figure S5. Scatter plots of significant spatial correlations between ECT-induced entropy alterations and neurotransmitter profiles.

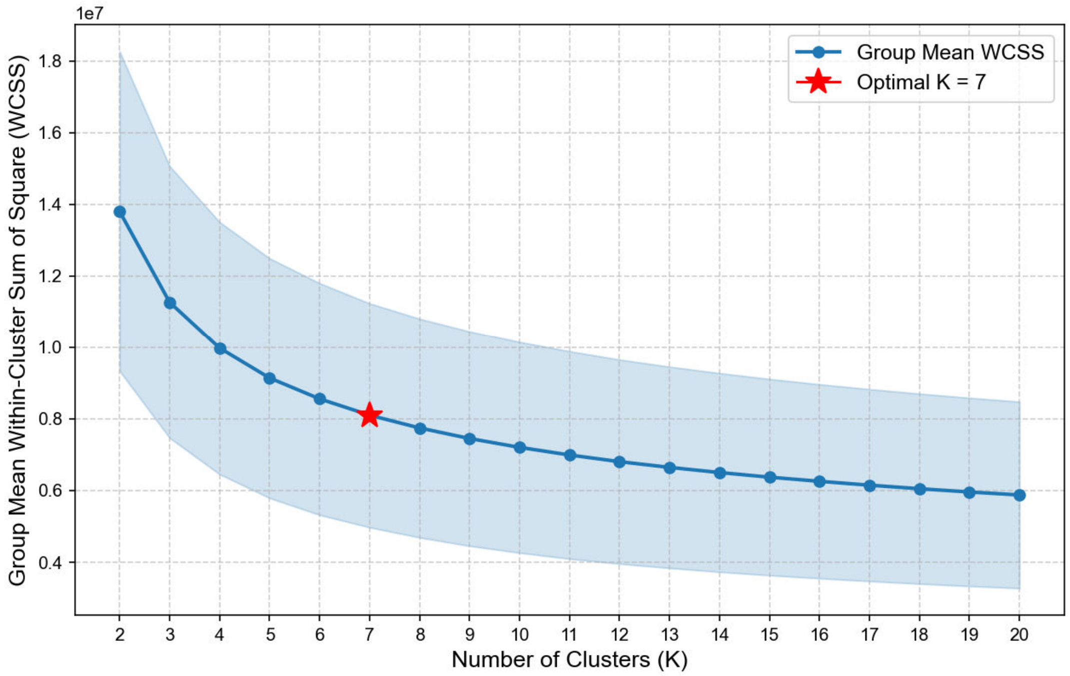


Figure S6. Group-level elbow curve. The blue line represents the group mean within-cluster sum of squares (WCSS) for K = 2 to 20 and the red star marks the optimal K value.
